# Transmission networks of SARS-CoV-2 in coastal Kenya during the first two waves: a retrospective genomic study

**DOI:** 10.1101/2021.07.01.21259583

**Authors:** Charles N. Agoti, Lynette Isabella Ochola-Oyier, Khadija Said Mohammed, Arnold W. Lambisia, Zaydah R. de Laurent, John M. Morobe, Maureen W. Mburu, Donwilliams O. Omuoyo, Edidah M. Ongera, Leonard Ndwiga, Eric Maitha, Benson Kitole, Thani Suleiman, Mohamed Mwakinangu, John Nyambu, John Otieno, Barke Salim, Jennifer Musyoki, Nickson Murunga, Edward Otieno, John Kiiru, Kadondi Kasera, Patrick Amoth, Mercy Mwangangi, Rashid Aman, Samson Kinyanjui, George Warimwe, My Phan, Ambrose Agweyu, Matthew Cotten, Edwine Barasa, Benjamin Tsofa, D. James Nokes, Philip Bejon, George Githinji

## Abstract

**Background:** The transmission networks of SARS-CoV-2 in sub-Saharan Africa remain poorly understood.

**Methods:** We undertook phylogenetic analysis of 747 SARS-CoV-2 positive samples collected across six counties in coastal Kenya during the first two waves (March 2020 - February 2021). Viral imports and exports from the region were inferred using ancestral state reconstruction (ASR) approach.

**Results:** The genomes were classified into 35 Pango lineages, six of which accounted for 79% of the sequenced infections: B.1 (49%), B.1.535 (11%), B.1.530 (6%), B.1.549 (4%), B.1.333 (4%) and B.1.1 (4%). Four identified lineages were Kenya specific. In a contemporaneous global subsample, 990 lineages were documented, 261 for Africa and 97 for Eastern Africa. ASR analysis identified >300 virus location transition events during the period, these comprising: 69 viral imports into Coastal Kenya; 93 viral exports from coastal Kenya; and 191 inter-county import/export events. Most international viral imports (58%) and exports (92%) occurred through Mombasa City, a key touristic and commercial Coastal Kenya center; and many occurred prior to June 2020, when stringent local COVID-19 restriction measures were enforced. After this period, local virus transmission dominated, and distinct local phylogenies were seen.

**Conclusions:** Our analysis supports moving control strategies from a focus on international travel to local transmission.

**Funding:** This work was funded by Wellcome (grant#: 220985) and the National Institute for Health Research (NIHR), project references: 17/63/and 16/136/33 using UK aid from the UK Government to support global health research, The UK Foreign, Commonwealth and Development Office.

## INTRODUCTION

By 31^st^ August 2021, at least 218 million cases of COVID-19 and > 4.5 million associated deaths were reported worldwide. By the same date, Kenya, an East Africa nation with a population of ∼50 million had reported 235,863 cases of SARS-CoV-2 and 4,726 associated deaths (MOH, 2021). Serological surveys indicated that Kenya’s COVID-19 epidemic had progressed further than could be discerned from the limited laboratory testing case reports that were available during the period (Etyang et al., 2021; Uyoga et al., 2021a). In the period after the peak of Kenya’s second wave (January-March 2021), nationwide anti-SARS-CoV-2 IgG prevalence based on analysis of blood donor samples was estimated to be 48.5% (Adetifa et al., 2021; Uyoga et al., 2021b).

In August 2021, 18 months after the first report of SARS-CoV-2 occurrence in Kenya, the country had experienced four major waves of SARS-CoV-2 infections (Brand et al., 2021). Despite this progression, the local SARS-CoV-2 spread patterns, like in many sub-Saharan Africa settings, remained poorly understood (Wilkinson et al.). The analysis presented here examined genome sequences from the first two waves in Coastal Kenya. Overall, the documented SARS-CoV-2 infections in the country during these early waves were concentrated in cities, especially Nairobi (∼42%), the capital, and Mombasa (∼8%), the country’s second largest city located on the Indian ocean coast (**Figure 1A-C**). In addition to Mombasa city, there are five other counties that make up Kenya’s Coastal region (Kilifi, Kwale, Taita Taveta, Tana River and Lamu) which has a total population size of ∼4.3 million.

**Figure 1.**
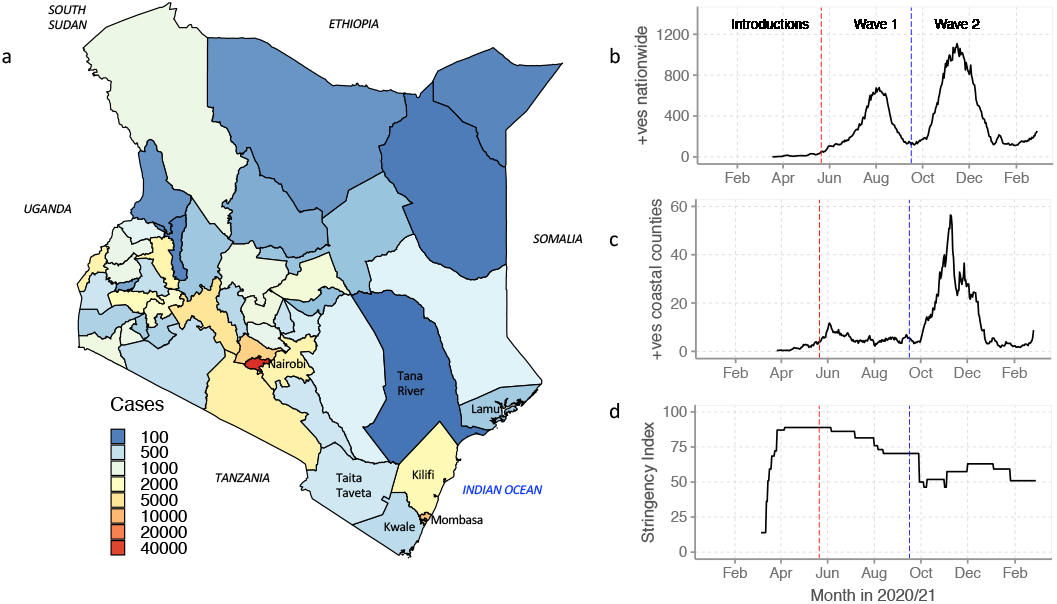
The SARS-CoV-2 epidemic in Kenya. Panel A, map showing administrative boundaries of the 47 counties in Kenya and the neighboring countries. The map is colored by the cumulative number of SARS-CoV-2 cases documented in each county as of 26^th^ February 2021. Panel B, reported daily new cases, 7-day-rolling average, in Kenya from March 2020 to February 2021 showing the first two national SARS-CoV-2 infection waves. Panel C. reported daily cases, 7-day-rolling average, across coastal Kenya during the study period. Panel D, Kenya government COVID-19 intervention levels during the study period as summarized by the Oxford stringency index (abbreviated SI, see details in materials and methods section).

Throughout the COVID-19 pandemic, genomic surveillance has been critical for tracking the spread of SARS-CoV-2 and investigating its evolutionary changes (Bugembe et al., 2020; Geoghegan et al., 2020; Oude Munnink et al., 2020; Worobey et al., 2020). In-country SARS-CoV-2 genome sequencing started in Kenya soon after the identification of the initial case and has remained a key tool informing public health response and tracking the progress of the local epidemic. We have previously reported on the genomic analysis of SARS-CoV-2 in Coastal Kenya during the early phase of the pandemic (up to July 2020) revealing introduction multiple lineages but with mainly Pango lineage B.1 establishing local transmission (Githinji et al., 2021). Here we investigated the lineage composition and diversity of the subsequent two waves (up to February 2021) and examine the patterns of further viral introductions, local evolution and spread of SARS-CoV-2 across the six Coastal counties of Kenya.

## RESULTS

### Infection waves in Coastal Kenya counties

The first cases of COVID-19 in Coastal Kenya were reported in Mombasa and Kilifi counties in March 2020. This was after initial detection in Nairobi City on 13^th^ March 2020. Following this, several COVID-19 related containment measures were put in place in March /April 2020 to curtail the spread of the virus including ban of movement into and out of infected areas. The national Oxford Stringency index in Kenya in April 2020 was at 89 (Brand et al., 2021), **Figure 1D**. However, two months later (June 2020), a steady removal of these restriction measures started leading to a SI of 75 by August 2020 and a SI <50 by October 2020. At the peak of the second wave, some of the restriction measures were reinstated increasing the SI to >=50, **Figure 1D**.

Mombasa county endured the earliest surge of SARS-CoV-2 cases in coastal Kenya with its first peak occurring in June 2020, **Figure 2A**. This was earlier than the first national peak that occurred in July-August of 2020. By 26^th^ February 2021 (the most recent date included in our analysis) three of the six coastal Kenya counties (i.e. Mombasa, Lamu and Taita Taveta) had experienced at least two major infection peaks while the other three (i.e. Kilifi, Kwale and Tana River) had experienced only a single peak. During this period covered, the Kenya Ministry of Health (MoH) reported 12,307 laboratory confirmed SARS-CoV-2 cases for all the six Coastal counties: Mombasa (n=8,450, 68.7%), Kilifi (n=2,458, 20.0%), Taita Taveta (n=567, 4.6%), Kwale (n=437, 3.6%), Lamu (n=309, 2.5%) and Tana River (n=85, 0.7%), **Figure 2A**.

**Figure 2.**
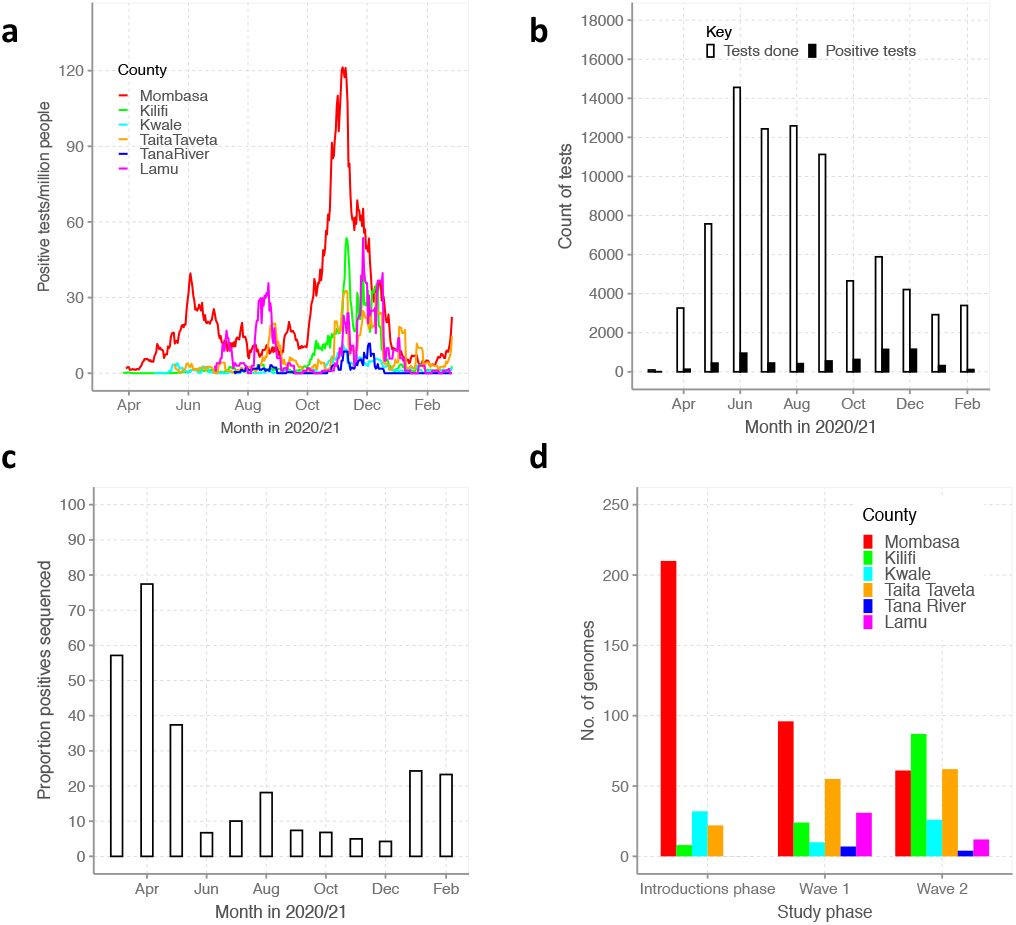
SARS-CoV-2 identified cases on the Kenyan Coast and testing and sequencing at KEMRI-Wellcome Trust Research Programme (KWTRP). Panel A, epidemic curves for each of the six Coastal Kenya counties derived from the daily positive case numbers, 7-day-rolling average, reported by the Ministry of Health. Panel B, monthly count of SARS-CoV-2 RT-PCR tests undertaken at the KWTRP and those positive during the study period. Panel C, monthly proportion of samples sequenced from total SARS-CoV-2 positives detected at KWTRP. Panel D, Distribution of the sequenced 747 samples by study period and across the 6 Coastal Kenya counties.

### Testing and sequencing at KWTRP

The flow of samples analyzed in this study is provided in **Supplementary-Figure 1**. Between 17^th^ March 2020 and 26^th^ February 2021, the KEMRI-Wellcome Trust Research Programme (KWTRP) tested at total of 82,716 nasopharyngeal/oropharyngeal (NP/OP) swab samples from the six Coastal counties. Of these, 6,307 (7.6%) were determined as SARS-CoV-2 positive (approximately half of the total reported by MoH), distributed by month as in **Figure 2B**. Overall, a total of 3,137 (6.8%) tests at KWTRP were positive from samples taken in Mombasa, 1,142 (8.8%) for Kilifi, 657 (4.5%) for Taita Taveta, 349 (12.7%) for Lamu, 437 (7.9%) for Kwale and 105 (12.0%) for Tana River.

Of the identified 3,137 RT-PCR positive samples, 747 (23.8%) were genome sequenced, distributed as follows; Mombasa: 367 (11.7%), Kilifi: 119 (10.4%), Taita Taveta: 139 (21.2%), Kwale: 68 (15.6%), Lamu: 43 (12.4%) and Tana River: 11 (10.5%), **Figure 2C**. The multiple sequence alignment of these coastal Kenya genomes and observed differences from the Wuhan-Hu-1 isolate is shown in **Supplementary-Figure 2**. The sequences were spread across the study period with 272 (36.4%) from the introduction phase (17^th^ March to 20^th^ May 2020 when nationally <50 new cases were reported per day), 223 (29.9%) from wave one (21^st^ May-15^th^ September 2020) and 252 (33.7%) from wave two (16^th^ September 2020 to 26^th^ February 2021), **Figure 1**. Overall, our sequencing rate corresponded to approximately one sequence for every 16 confirmed cases.

Most of the sequenced isolates were from males (n=443, 59.3%), persons aged between 30-60 years (n=420, 56.2%) and persons of Kenyan nationality (n=509, 68.1%), **Table 1**. The second largest nationality of the individuals providing sequenced samples was Tanzania (n=34) followed by South Africa (n=5). A total of 110 samples (14.7%) had been collected from persons with a recent international travel history (<14 days) but a considerable proportion of the sequenced samples were obtained from persons whose travel history information was missing (n=373, 49.9%).

**Table 1:**
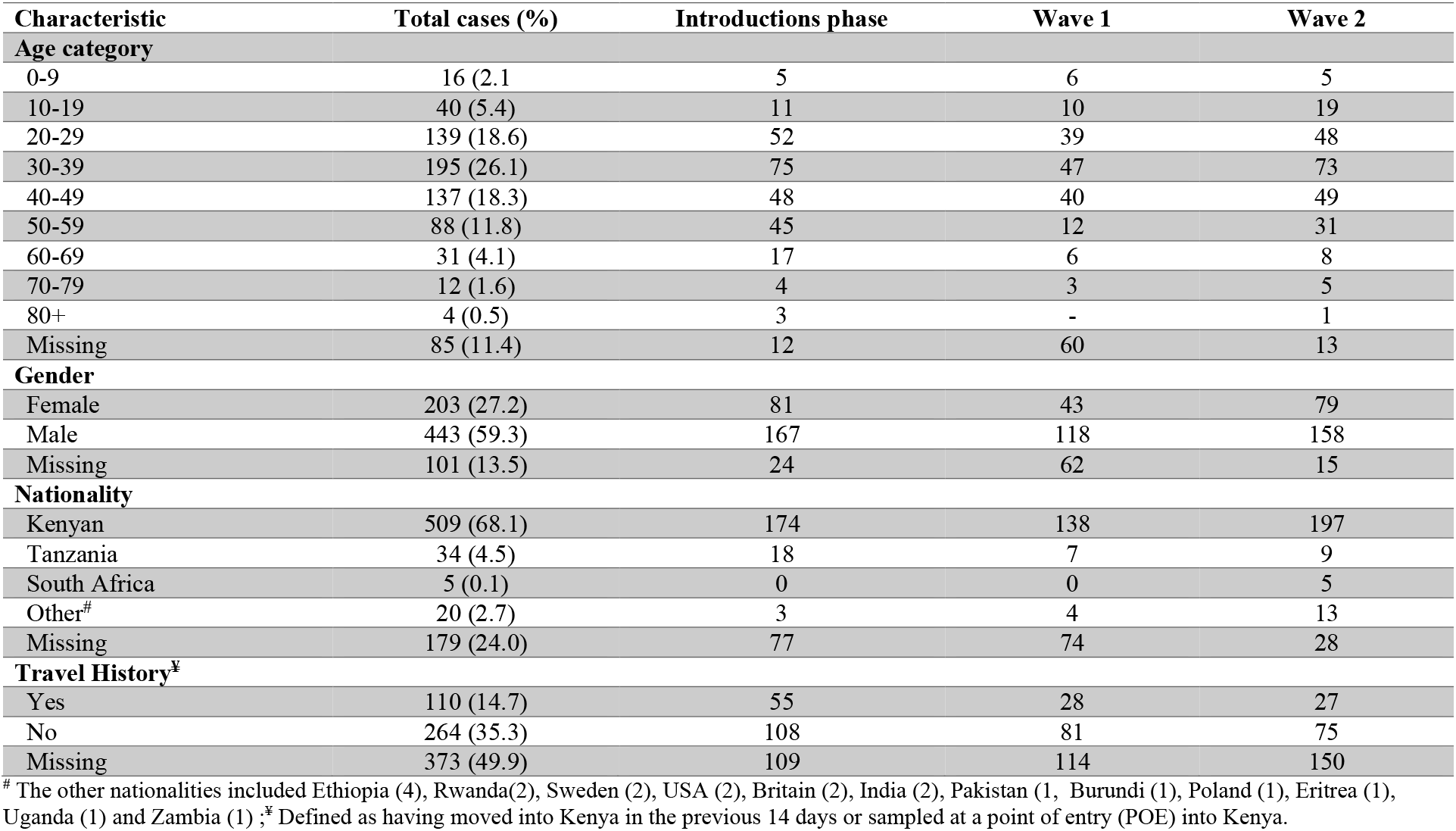
Demographics of the sequenced SARS-CoV-2 cases in coastal Kenya

### Lineage dynamics in Coastal Kenya

As a reference, we adopted the PANGOLIN (<u>P</u>hylogenetic <u>A</u>ssignment of <u>N</u>amed <u>G</u>lobal <u>LIN</u>eages) lineage nomenclature (Rambaut et al., 2020). The 747 genomes were classified into 35 Pango lineages, including four first identified in Kenya (N.8, B.1.530, B.1.549 and B.1.596.1), **Table 2 and Figure 3A**. During the introductions phase we detected 14 lineages while during wave one we detected 21 lineages (10 of which were being identified locally for the first time) and during wave two we detected 19 lineages (seven of which were being identified locally for the first time), **Figure 3B**. Seventeen lineages (48.6%) were identified in three or more samples with the top six lineages accounting for 78.8% of the sequenced infections; B.1 (n=369, 49.4%, European lineage which predominated in the Northern Italy outbreak early in 2020), B.1.535 (n=82, Australian lineage), B.1.530 (n=42, 5.6%, Kenyan lineage), B.1.530 (n=42, 6%, Kenyan lineage), B.1.549 (n=33, 4.4%, Kenyan lineage), B.1.333 (n=32, 4.3%, Washington USA lineage), B.1.1 (n=31, 4.1%, European lineage).

**Table 2.** Lineages observed in coastal Kenya, their county distribution, global history and VOC/VOI status

| Lineage | Frequency (%) | Mombasa | Kilifi | Kwale | Taita Taveta | Tana River | Lamu | Earliest date | Number assigned | Description |
| --- | --- | --- | --- | --- | --- | --- | --- | --- | --- | --- |
| **A** | 22 ( 2.9 ) | 4 | - | 13 | 5 | - | - | 2019-12-30 | 2164 | Root of the pandemic lies within lineage A. Many sequences originating from China |
| **A.23** | 2 ( 0.3 ) | - | 1 | 1 | - | - | - | 2020-06-08 | 99 | Ugandan lineage |
| **A.23.1** | 6 ( 0.8 ) | 2 | 1 | 1 | 2 | - | - | 2020-10-21 | 1109 | International lineage with a number of variants of potential biological concern, including 681R |
| **A.25** | 2 ( 0.3 ) | 2 | - | - | - | - | - | 2020-06-08 | 46 | Ugandan lineage |
| **AL.1** | 4 ( 0.5 ) | 3 | - | - | 1 | - | - | 2020-02-23 | 1268 | Alias of B.1.1.231.1, Canada lineage |
| **B** | 11 ( 1.5 ) | 10 | 1 | - | - | - | - | 2019-12-24 | 5408 | Second major haplotype (and first to be discovered) |
| **B.1** | 369 ( 49.4 ) | 201 | 65 | 19 | 65 | 8 | 11 | 2020-01-24 | 71452 | A large European lineage the origin of which roughly corresponds to the Northern Italian outbreak early in 2020. |
| **B.1.1** | 31 ( 4.1 ) | 12 | 3 | 4 | 12 | - | - | 2020-02-16 | 42839 | European lineage with 3 clear SNPs `28881GA`, `28882GA`, `28883GC` |
| **B.1.1.1** | 4 ( 0.5 ) | 1 | 1 | 2 | - | - | - | 2020-03-02 | 2796 | England |
| **B.1.1.33** | 13 ( 1.7 ) | 2 | 1 | - | - | - | 10 | 2020-03-01 | 2039 | Brazilian |
| **B.1.1.7** | 2 ( 0.3 ) | 2 | - | - | - | - | - | 2020-02-07 | 986233 | UK lineage of concern, associated with the N501Y mutation. |
| **B.1.13** | 1 ( 0.1 ) | 1 | - | - | - | - | - | 2020-03-09 | 398 | UK lineage |
| **B.1.153** | 1 ( 0.1 ) | - | - | 1 | - | - | - | 2020-03-01 | 773 | English lineage with some Australian, New Zealand and European sequences |
| **B.1.157** | 2 ( 0.3 ) | 2 | - | - | - | - | - | 2020-02-03 | 324 | UK/ Spanish lineage |
| **B.1.160** | 2 ( 0.3 ) | - | - | 1 | - | - | 1 | 2020-02-06 | 24207 | Large European lineage, sequences from BeNeLuX, Denmark, Switzerland, Hungary and UK. |
| **B.1.177** | 1 ( 0.1 ) | - | 1 | - | - | - | - | 2020-02-14 | 69319 | Major mostly European lineage, result of opening borders summer 2020 |
| **B.1.179** | 2 ( 0.3 ) | 2 | - | - | - | - | - | 2020-03-09 | 231 | Denmark |
| **B.1.222** | 2 ( 0.3 ) | 2 | - | - | - | - | - | 2020-02-24 | 562 | Scottish lineage |
| **B.1.229** | 2 ( 0.3 ) | 2 | - | - | - | - | - | 2020-03-19 | 104 | UK lineage |
| **B.1.281** | 1 ( 0.1 ) | 1 | - | - | - | - | - | 2020-04-08 | 41 | Bahrain lineage |
| **B.1.333** | 32 ( 4.3 ) | 27 | 2 | 3 | - | - | - | 2020-03-03 | 188 | USA lineage WA |
| **B.1.351** | 25 ( 3.3 ) | 6 | 4 | 8 | 7 | - | - | 2020-03-27 | 25467 | Lineage of concern detected in South Africa |
| **B.1.393** | 1 ( 0.1 ) | - | 1 | - | - | - | - | 2020-05-29 | 37 | Uganda |
| **B.1.525** | 1 ( 0.1 ) | - | 1 | - | - | - | - | 2020-12-11 | 6729 | International lineage with E484K, del69-70 among other defining mutations. |
| **B.1.530** | 42 ( 5.6 ) | 8 | 7 | - | 26 | - | 1 | 2020-10-20 | 54 | Kenya |
| **B.1.535** | 82 ( 11.0 ) | 50 | 11 | 10 | 10 | 1 | - | 2020-02-03 | 2573 | Australia |
| **B.1.549** | 33 ( 4.4 ) | 11 | 17 | 3 | 2 | - | - | 2020-07-17 | 51 | Kenya + England |
| **B.1.551** | 2 ( 0.3 ) | - | - | - | 2 | - | - | 2020-07-31 | 751 | USA lineage |
| **B.1.558** | 1 ( 0.1 ) | 1 | - | - | - | - | - | 2020-04-06 | 190 | USA/ Mexico lineage |
| **B.1.593** | 2 ( 0.3 ) | - | - | - | - | 2 | - | 2020-08-27 | 90 | USA lineage |
| **B.1.596.1** | 15 ( 2.0 ) | 10 | 2 | 2 | 1 | - | - | 2020-09-07 | 54 | Kenyan lineage |
| **B.1.612** | 7 ( 0.9 ) | - | - | - | 6 | - | 1 | 2020-03-11 | 380 | USA lineage, split out from B.1.361 |
| **B.4** | 3 ( 0.4 ) | 3 | - | - | - | - | - | 2020-01-18 | 376 | Iranian lineage and global exports |
| **B.4.7** | 1 ( 0.1 ) | 1 | - | - | - | - | - | 2020-03-14 | 68 | Africa and UAE |
| **N.8** | 20 ( 2.7 ) | 1 | - | - | - | - | 19 | 2020-06-23 | 14 | Alias of B.1.1.33.8, Kenyan |

**Figure 3.**
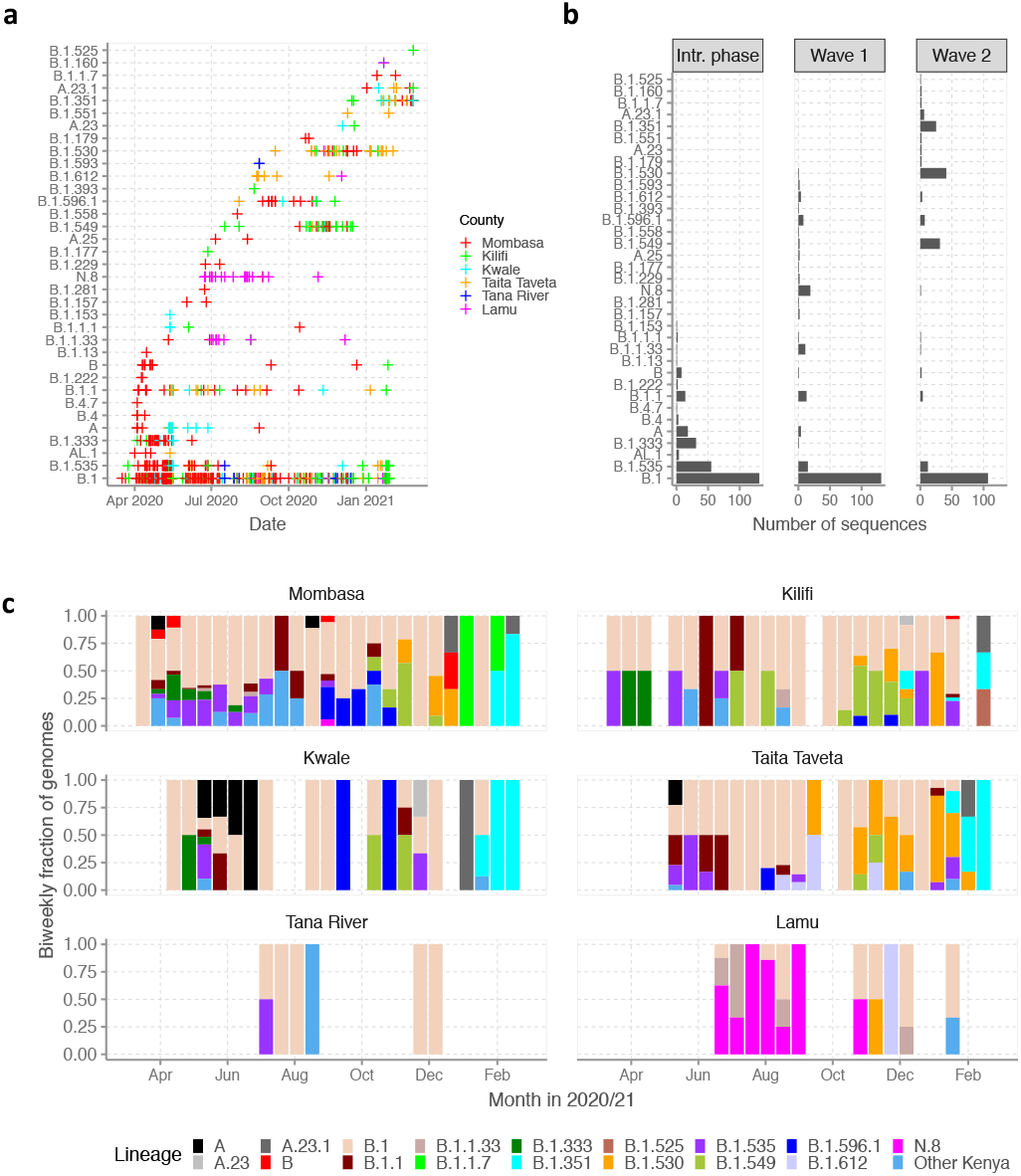
Lineage introductions and temporal dynamics in Coastal Kenya. Panel A, SARS-CoV-2 Pango lineages in the sequenced 747 coastal Kenya samples and timing of detections. Panel B, Lineage detections by study phase. Intr. phase refers to introduction phase. Panel C, Biweekly distribution of the common lineages identified across the six counties presented as proportions. Lineages detected in less than three cases were put together and referred to as “other lineages”. This group included 18 lineages namely: A.25, AL.1, B.1.1.1, B.1.13, B.1.153, B.1.157, B.1.160, B.1.177, B.1.179, B.1.222, B.1.229, B.1.281, B.1.393, B.1.551, B.1.558, B.1.593, B.4 and B.4.7.

Lineage B.1 was the first to be detected in the region and comprised the initial cases identified in Mombasa, Kilifi, Kwale and Tana River, **Figure 3A and C**. This lineage comprised 48.2% (n=131) of the viruses we sequenced from the introduction phase, 58.7% (n=131) from wave one and 42.5% (n=107) from wave two. Overall, B.1 accounted for most of the virus sequences detected in all counties except Lamu where lineages B.1.1.33 and N.8 (alias lineage B.1.1.33.8) were most frequent (together accounting for 67.5% of Lamu cases, especially wave one, first detected there on 23^rd^ June 2020), **Table 2**. Only three additional sequenced cases of lineage B.1.1.33 occurred outside Lamu County (two Mombasa and one in Kilifi) and only one additional sequenced case of lineage N.8 occurred outside Lamu County (in Mombasa).

During wave one, 18 cases of lineage A were observed. Most (56%) of these were identified in Kwale county (one of the Kenya counties bordering Tanzania, **Figure 1A**), and the lineage comprised 19% of all cases identified Kwale county, **Figure 3C**. Only two other counties observed lineage A cases: Mombasa (n=4) and Taita Taveta (n=5). Lineage B.1.535, which was the second most common lineage in the region was mostly observed in during wave one and predominantly in Mombasa County with limited number of cases observed in Kilifi, Kwale and Taita Taveta, **Table 2**.

During the wave two period, there was an emergence of three lineages first identified in Kenya: (a) lineage B.1.549 (first observed 17^th^ July 2020) which was responsible for a significant number of the sequenced infections in Kilifi (n=17), Mombasa (n=11), Taita Taveta (n=2), and Kwale (n=3), (b) lineage B.1.596.1 (first observed 03^rd^ August 2020) and detected in Mombasa, Kilifi, Kwale and Taita-Taveta and (c) lineage B.1.530 (first observed 15^th^ September 2020) that predominated infections in Taita Taveta (n=26) with a fewer cases observed in Mombasa (n=8), Kilifi (n=7) and Lamu (n=1), **Figure 3A** and **B**.

Two variants of concern (VOC) were detected during the study period: Beta (i.e. lineage B.1.351 in 25 samples); and Alpha (i.e lineage B.1.1.7 in two samples (Konings et al., 2021)). The earliest B.1.351 infections were identified in international travelers from South Africa in mid-December 2020 while the earliest detected B.1.1.7 case was in second week of January 2020 in a local with no history of recent international travel. We also detected two cases of A.23 and six cases of A.23.1 after the peak of wave two (both lineages first identified in Uganda, and A.23.1 was considered a variant of interest (VOI) in the region in early 2021 (Bugembe et al., 2021a)).

### Lineage dynamics with a widening scale of observation

We compared the temporal prevalence patterns of the lineages identified in Coastal Kenya to Eastern Africa, Africa and a global sub-sample, **Figure 4**. In these comparative genome sets, we identified 97 Pango lineages for Eastern Africa, 261 for Africa and 990 for the global sub-sample. Note that by May 2021, a total of 1,293 Pango lineages has been assigned worldwide since the beginning of the pandemic (O’Toole et al., 2021) indicating very high representativeness in our global subsample.

**Figure 4.**
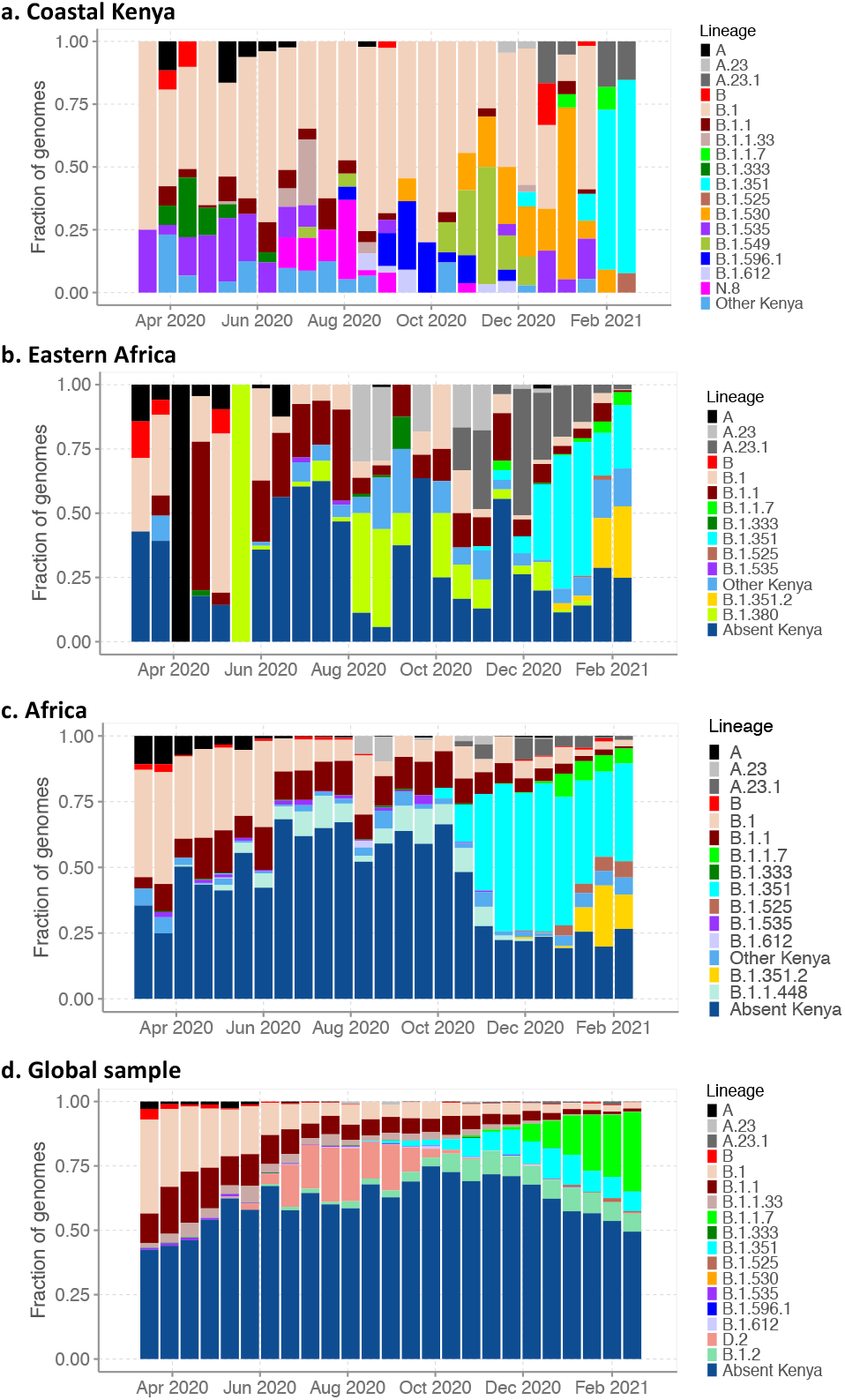
Lineage dynamics at widening scales of observation. Panel A, Biweekly distribution of detected lineages in Coastal Kenya from the sequenced 747 genomes. Panel B, biweekly distribution of detected lineages in Eastern Africa from 2,261 contemporaneous genomes from 10 countries whose contemporaneous data were available in GISAID. Lineages not observed in the Coastal Kenya dataset have been lumped together as “Absent Kenya” but with the those appearing among top six and not observed in Kenya indicated just below the other Kenya group. Panel C, weekly distribution of detected lineages in Africa from 12,191 contemporaneous genomes from 39 countries that were available in GISAID. Panel D, weekly distribution of detected lineages in a global sub-sample of 25,511 contemporaneous genomes from 139 countries that were compiled from GISAID (see detail in methods section).

In our Eastern Africa sample (n=2,261), the top lineages were also observed in the Coastal Kenya data (B.1.351, A.23.1, B.1.1 and B.1) except lineage B.1.351.2 (n=180, sub-lineage of B.1.351 circulating in Mayotte) and B.1.380 (n=128, a Rwanda lineage (Butera et al., 2021)), **Figure 4B**. In the Africa sample (n=12,191), four of the six top lineages (B.1, B.1.1, B.1.351 and B.1.1.7) were also identified in Coastal Kenya. The undetected two lineages were B.1.1.448 (n=392) mainly observed South Africa samples, and B.1.351.2, **Figure 4C**. In the global sub-sample (n=24,511) four of the top six lineages (B.1, B.1.1, B.1.1.7 and B.1.351) were observed in the samples from Coastal Kenya. The unobserved lineages were D.2 (Alias of B.1.1.25.2, an Australian lineage) and B.1.2 (a USA lineage), **Figure 4D**.

At the time of the second wave in Kenya, the Alpha and Beta VOC were already widespread across Eastern Africa and Africa but there were only sporadic detections from our Coastal Kenya dataset. Although in the comparison data lineage B.1 occurred in substantial proportions across the different scales early in the pandemic, its prevalence diminished faster overtime in the outside Coastal Kenya sample sets when compared to the Coastal Kenya dataset set. Greater than 95% of the lineages comprising infections globally were not seen in the Coastal Kenya samples.

### SARS-CoV-2 diversity in Coastal Kenya

We constructed both a mutational maximum likelihood (ML) (i.e. non-time resolved) and a time-resolved phylogeny of the Coastal Kenya genomes both while including a random global reference set of genomes (n=4000) shown in **Supplementary-Figure 3** and **Figure 5A**, respectively. These analysis showed that (i) the Coastal Kenya genomes were represented across many but not all of the major phylogenetic clusters observed globally, ii) some of the coastal Kenya clusters expanded after introduction whereas others did not (observed as singletons) and iii) all counties appeared to have had multiple lineage/variant introductions with some clusters comprising genomes detected across multiple counties, **Figure 5A**. There was considerable correlation of the root-to-tip genetic distance and the date of sampling dates (r^2^=0.49), **Figure 5B**.

**Figure 5.**
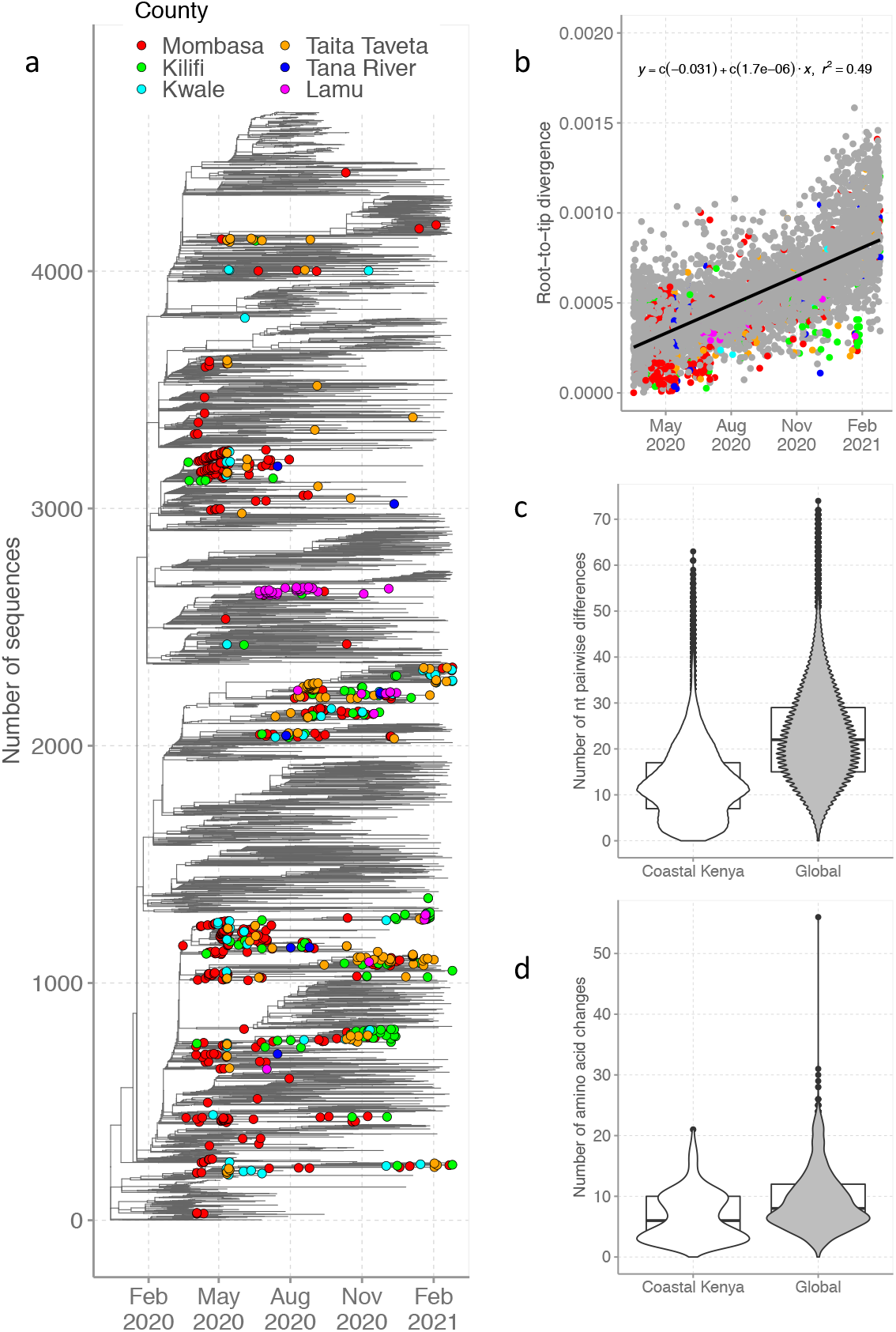
SARS-CoV-2 diversity in Coastal Kenya. Panel A, Time-resolved global phylogeny that combined 732 Coastal Kenya SARS-CoV-2 genomes and 3,958 global reference sequences (after removal of sequences not fitting molecular clock-like evolution using Tree-Time). Panel B, Root-to-tip regression analysis coastal Kenya (colored by county) and global (shown in grey) sequences with determination of correlation co-efficient (r^2^). Panel C, comparison of pairwise nucleotide differences of the 732 coastal Kenya genomes included on the phylogenetic tree and the 3,958 global genomes. Panel D, comparison of the number of amino acid substitutions of the coastal Kenya genomes versus the global dataset relative to the Wuhan-Hu-1 reference genome.

The coastal Kenya genomes had a fewer pairwise nucleotide differences (median 12, interquartile range (IQR): 7-17) compared to the global subsampled comparison dataset median 22, IQR: 15-29), **Figure 5C**. A similar difference was observed at amino acid level, with the coastal Kenya genomes showing a median of six amino acid substitutions (IQR: 3-10) while the global dataset showed a median of eight amino acid substitutions (IQR: 6-12), **Figure 5D**.

For detailed investigation into the local SARS-CoV-2 genetic diversity, we reconstructed time-resolved lineage specific phylogenetic trees are shown in **Figure 6**. The three lineages first identified in Kenya (B.1.530, B.1.549 and B.1.596.1) were found to (a) possess significant diversity consistent with widescale spread within Kenya (**Figure 6C-E**), (b) formed multiple county-specific sub-clusters and (c) show local sequences interspersed with global comparison genomes from the same lineage implying potentially export (or import) events, **Figure 6D**. However, the picture painted by lineage N.8 was different. This lineage was mainly detected in Lamu forming a single monophyletic group (**Figure 6H**) when co-analyzed with its precursor lineage B.1.1.33, an observation consistent with a single introduction as B.1.1.33 then local evolution expansion.

**Figure 6.**
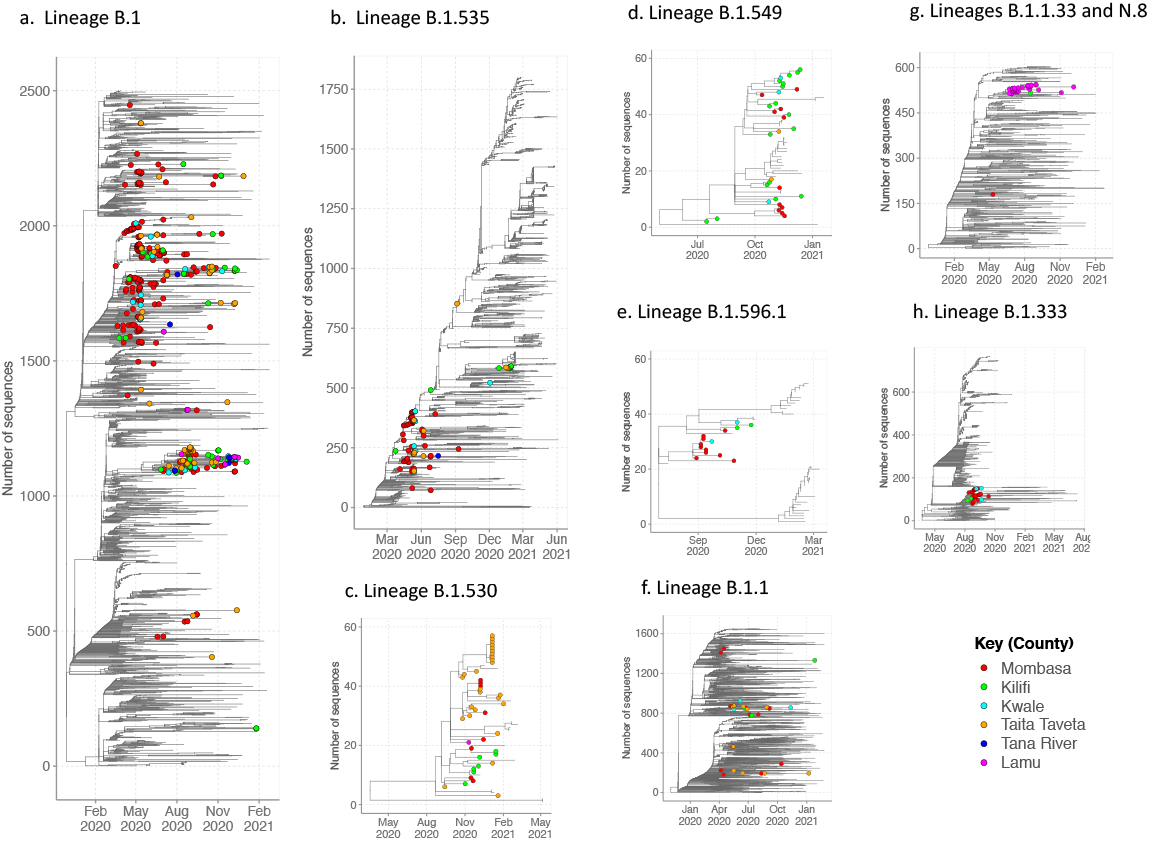
Time-resolved lineage-specific phylogenetic trees for the commonly detected lineages in Coastal Kenya. The Coastal Kenya genomes are indicated with filled circles colored by county. Panel A, phylogeny for lineage B.1 that combined 369 Coastal Kenya genomes and 2,319 global reference sequences. Panel B, phylogeny for lineage B.1.535 that combined 82 Coastal Kenya genomes and 1,867 global sequences belonging also to B.1.535. Panel C, phylogeny for lineage B.1.530 that combined 42 Coastal Kenya genomes and 15 global sequences belonging also to B.1.530. Panel D, phylogeny for lineage B.1.549 that combined 33 Coastal Kenya genomes and 23 sequences from other countries of lineage B.1. 549. Panel E, phylogeny for lineage B.1.596.1 combines 15 Coastal Kenya genomes and 36 global reference sequences. Panel F, phylogeny for lineage B.1.1 that combined 31 Coastal Kenya genomes and 1,629 global sequences belonging to lineage B.1.1. Panel G, phylogeny for lineage N.8 that combined 20 Coastal Kenya genomes of lineage N.8, 13 Coastal Kenya genomes of lineage B.1.1.33 and 570 global sequences of lineage B.1.1.33. Panel H, phylogeny for lineage B.1.333 combines 32 Coastal Kenya genomes and 753 global reference sequences.

In **Figure 7**, we present a comparison of both the number of nucleotide and amino acid substitutions for each major lineage between the coastal Kenya genomes and the global subsample. The divergence of the lineages from the original Wuhan sequence at the global level was closely mirrored at the local coastal Kenya level. The most divergent lineages among the subset we examined were three of the Kenya specific lineages (B.1.530, B.1.549 and B.1.596.1). The least genetically divergence lineages from the original Wuhan SARS-CoV-2 strain were viruses occurring within lineage B.1.333 and lineage B.1.535, **Figure 7**.

**Figure 7.**
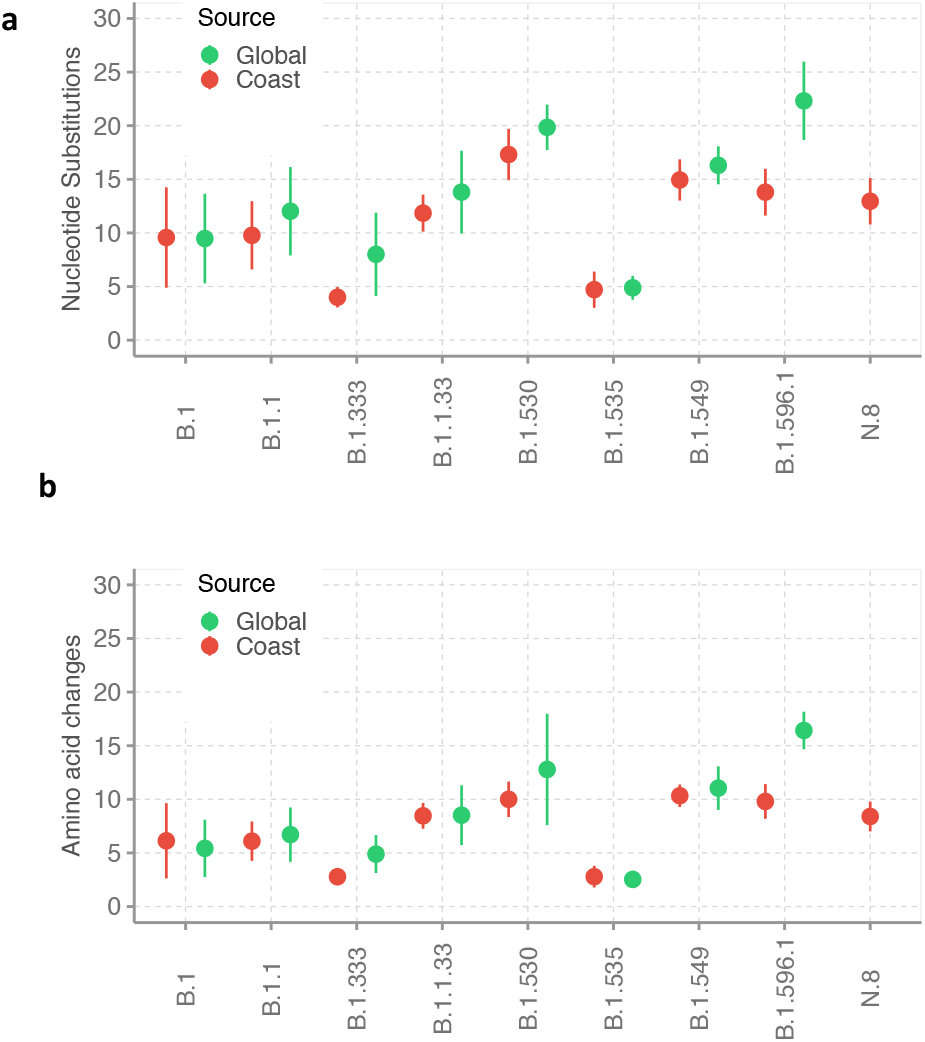
Level of genetic divergence of the major detected lineages in coastal Kenya. The sequences were first aligned to the 2019 Wuhan-Hu-1 isolate and the number of changes compared to isolates across the globe of the same lineage. Panel A shows the divergence at nucleotide sequence level. Panel B shows the divergence at amino acid sequence level focusing on synonymous substitutions.

### Viral imports and export from Coastal Kenya

We used ancestral location state reconstruction of the dated phylogeny **(Figure 5A)** to infer the number of viral import and exports (Sagulenko et al., 2018). In total, between March 2020 and February 2021, we detected 357 location transition events within and between the six counties and the outside world. Sixty-nine of these were viral import events into Coastal Kenya from outside and 93 were viral exports events from Coastal Kenya to outside populations, **Figure 8A-C**. A total of 191 import/export events were detected between the six Coastal counties.

**Figure 8.**
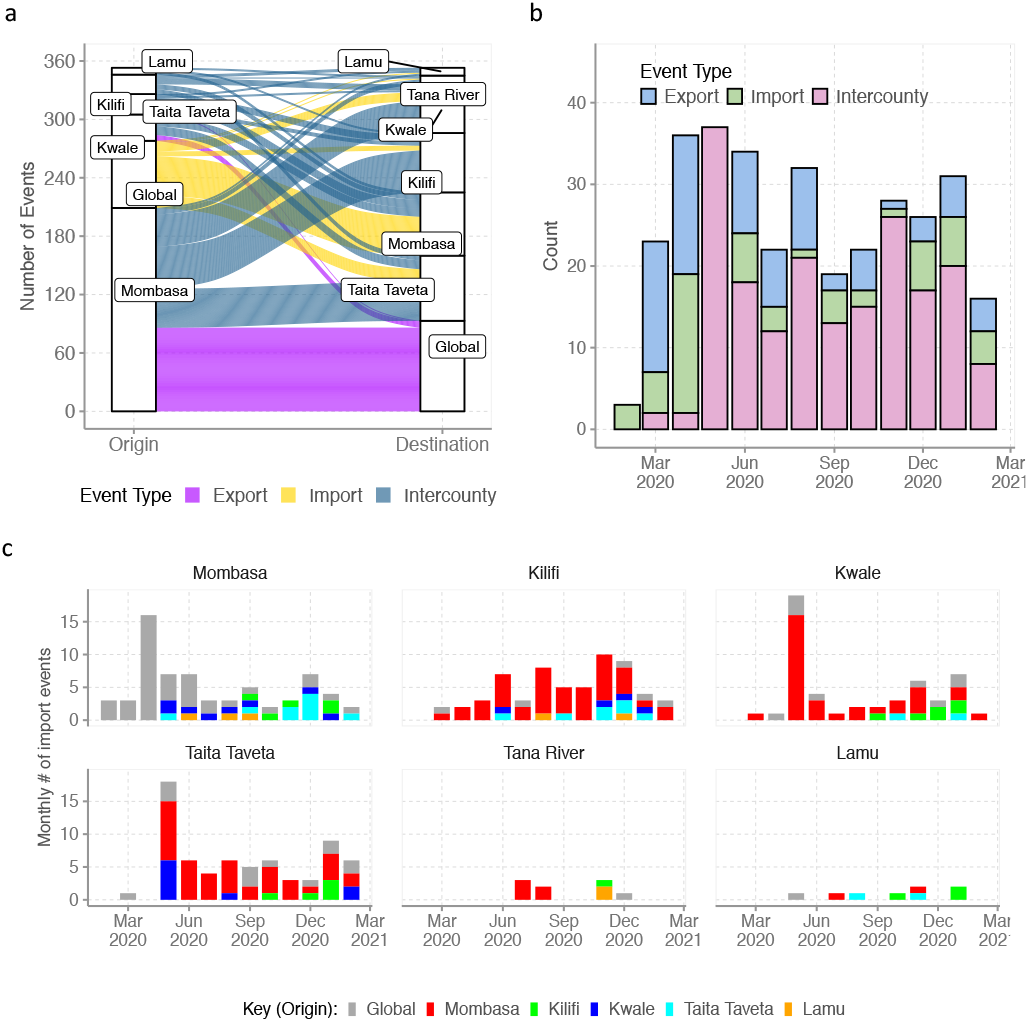
Viral imports and export from Coastal Kenya. Panel A, an alluvium plot showing the number of imports and exports of viral lineages from Coastal Kenya shown as an alluvium plot. Panel B, monthly location transition events observed in the Coastal Kenya and global genomes shown as either viral export, import or inter-county transmission. Panel C, monthly number of import events into the individual Coastal counties and their origins.

Virus imports into the region occurred through Mombasa (n=40, 60%), Kwale (n=9, 13%), Taita Taveta (n=13, 19%), Kilifi (n=5, 7%) and Lamu (n=1, 1%) while virus export from the region occurred through Mombasa (n=86, 41%), Kwale (n=6, 6%) and Taita Taveta (n=1, 1%). Of the 69 detected viral imports, 34 (49%) occurred during the introductions phase, 16 (23%) occurred during wave one and 19 (28%) during wave two while of the 93 detected viral exports, 43 (46%) occurred during introductions phase, 31 (33%) during wave one and 19 (20%) during wave two, **Figure 8B and 8D**.

The analysis revealed the top six country sources of importation into coastal Kenya as; USA, (n=19, 31%), England (n=10, 16%), Uganda (n=6, 10%), Japan (n=5, 8%) and Mozambique (n=3, 5%), **Supplementary-Figure 4 and Supplementary-Figure 5**. On the other hand, the top six country destinations of viral export from coastal Kenya were USA (n=22, 17%), Australia (n=16, 13%), India (n=14, 11%), Uganda (n=8, 6%), England (n=7, 6%) and Scotland (n=4, 3%).

To infer the sensitivity of our analysis, we repeated the import/export analysis using a (a) different global sub-sample (**Supplementary-Figure 4)** and (b) with a normalized subsample of the coastal Kenya genomes accounting for total reported infections per county, **Supplementary-Figure 5**. The reanalysis came to roughly similar conclusions as above and the results are presented in **Supplementary-Table 1**.

### Amino acid changes in the Coastal Kenya genomes

We scanned the entire SARS-CoV-2 genome for amino acid changes from the Wuhan reference genome in the 747 Coastal Kenya sequences. Below we summarize changes that were identified in ≥5 genomes. We observed 20 changes in the spike (S) protein meeting the set criterion, 15 in the nucleocapsid (N) protein, 14 in ORF3a, 13 in ORF1b, nine in the ORF1a, seven in ORF8, five in ORF6, and one in the envelope (E) protein. No codon changes, meeting of criterion was observed in the matrix (M) protein, ORF7a, ORF7b or ORF10. The most common amino acid changes observed in the Coastal Kenya genomes were S: D614G (94.4%), ORF1b: P314L (92.9%), ORF3a: Q57H (15.5%), ORF1a: T265I (15.4%) and N: R195K (12.9%), **Supplementary-Figure 6**. Several lineage-defining mutations were observed for the Kenya-specific lineages as summarized in **Supplementary-Figure 7**.

## DISCUSSION

We present evidence of continuous additional introductions of SARS-CoV-2 into Coastal Kenya during the first year of the pandemic documenting at least 69 independent viral introduction events into the region. Multiple introductions were observed even at county level, with extensive inter-county spread observed after the early phase period. Strikingly, many of the imports and exports from the region occurred through the city of Mombasa, a major commercial, industrial and tourist destination in the region. This observation highlights Mombasa’s central role as a gateway for virus entry into coastal Kenya and the significance continuous surveillance in this county for early detection of variants in coastal Kenya populations.

During the early phase of the pandemic several Pango lineages were introduced into the region (A, B, AL.1, B.1, B.1.1, B.1.1.1, B.1.1.33, B.1.13, B.1.153, b.1.222, B.1.333, B.1.535, B.4 and B.4.7). However, it is lineage B.1 which has a European origin, that became predominant, 48.2% of the cases (Githinji et al., 2020). This lineage had the D614G change in the spike protein which has been found to enhance viral fitness (Baric, 2020) and this may have boosted its local transmission and early dominance. Notably also the import/export analysis found that the majority of viral imports to coastal Kenya had an Origin in USA, England and neighboring Uganda. Viral exports from costal Kenya happened to USA, Uganda, Australia among other destinations.

During the national wave one, the Coastal region observed SARS-CoV-2 transmission mainly in counties of Mombasa, Taita Taveta and Lamu. The key lineages observed then were B.1 (58.7%), N.8 (8.517%), B.1.535 (6.7%), B.1.1 (5.8%). Notably, lineage N.8 was specific to Lamu County. Lineage N.8 has not been observed outside Kenya and its precursor lineage (B.1.1.33) was observed earlier in South America, mainly Brazil. Lineage N.8 may have arisen from an introduction event of B.1.1.33 into Lamu county, and then local evolution and localization to Lamu. The N.8 lineage has seven characteristic lineage defining mutations including S: D614G and R203K, G204R and I292T on the nucleocapsid protein.

The national wave two started in October 2020 and led to significant viral transmission in all the six coastal counties. In Kilifi, Tana River and Kwale counties, this was the first significant peak. This wave two was majorly comprised mainly three lineages: B.1 (42%), B.1.530 (16%), and B.1.549 (12%). Lineages B.1.530 and B.1.549, first identified in Kenya, and may have arisen from local evolution of lineage B.1. Lineage B.1.530 has six characteristic mutations including spike P681H change adjacent to the biologically important furin cleavage site while lineage B.1.549 has seven characteristic mutations, five occurring in the ORF1a or ORF1b. Both two lineages have now been observed in a few other countries albeit in small numbers: seven countries for B.1.530 (Rwanda, Netherlands, Germany, Denmark, USA, Japan and Australia) and three countries for B.1.549 (England, USA, Canada).

Near the tail end of the national wave two, we detected cases of the Beta VOC (B.1.351) and the Alpha VOC (B.1.1.7). Beta was first to be observed, initially in Kilifi in two asymptomatic international travelers in mid-December 2020. Alpha was subsequently detected in a local who presented to a Mombasa clinic in the second week of January 2021. In the subsequent weeks up to the end of the period covered by this analysis (February 2021), only one additional Alpha case was detected in our sequenced samples unlike lineage Beta which continued to be detected sporadically in January and February 2021 especially in the counties of Taita Taveta and Kwale which border Tanzania.

When we compared the local lineage patterns to the global patterns, it was notable that although several lineages were detected globally (>900), only a small fraction (<10%) of this were documented locally (O’Toole et al., 2021). The VOC were already extensively spread across Eastern Africa (Beta VOC), Africa (Beta VOC) and worldwide (Alpha VOC) in the last quarter of 2020 unlike the findings here for costal Kenya. This implies that a slight lag occurred in the VOC arrival and large-scale spared in Coastal Kenya perhaps due to remoteness and public health measures that have remained in place during the period.

While Mombasa, Taita Taveta and Lamu counties had experienced two waves of infections by February 2021, Kilifi, Tana River and Kwale counties had experienced only a single major wave. This may have been brought about by differences in population size, density, and accessibility for these different counties. For instance, Mombasa is densely populated (∼5,604.64 persons/sq.km, 2019 census), has a large seaport and an international airport while Lamu has a sparsely populated mainland and with several islands (total population 143,920, population density, 21.84 persons/sq.km). Taita Taveta and Kwale counties are primarily rural counties bordering Tanzania which had a largely uncontrolled SARS-CoV-2 epidemic during the study period (Rice et al., 2021). Tana River is the most remote and sparsely populated (8.9 persons/sq.km) county in the region.

Our study contributes to the limited but growing literature illuminating SARS-CoV-2 transmission patterns in sub-Saharan Africa (Bugembe et al., 2021a; Bugembe et al., 2021b; Tegally et al., 2021a; Tegally et al., 2021b; Wilkinson et al.). The patterns revealed here inform on frequency, sources and routes of introductions of SARS-CoV-2 into Coastal Kenya and have implications on future control strategies. The key limitations of our analysis include, first, the samples we sequenced were only those available to us through the rapid response teams (RRTs) whose case identification protocols were altered at the different study pandemic phases. Second, we only sequenced ∼20% of the positive samples identified in our laboratory, the majority from the introduction phase and were prioritizing samples with a Ct value of <30.0. Third, the sampling across the six Coastal counties was not uniform, probably in part due to varied distance from our testing centre located in Kilifi County.

In conclusion, we show that the first two SARS-CoV-2 waves in Coastal Kenya observed transmission of both newly introduced and potentially locally evolved variants. New variant introductions were mainly through Mombasa city and originated USA, England, Uganda among other predicated sources. Only a limited number of the many introduced variants progressed to transmit extensively. Unlike in the global contemporaneous sample, we did not find evidence of extensive local transmission of the global VOC during wave two. Thus, we infer that it is more likely that the relaxation of some of the interventions (e.g., reopening of learning institutions, airspace, bars and restaurants) that drove the second wave of infections. Our study shows the importance of detailed local genomic surveillance even in remote and under-resourced settings to understand spread patterns of SARS-CoV-2 to optimize local interventions.

## MATERIALS AND METHODS

### Ethical statement

Samples analysed here were collected under the MoH protocols as part of the national response to the COVID-19 pandemic. The whole genome sequencing study protocol was reviewed and approved by the Scientific and Ethics Review Committee (SERU), Kenya Medical Research Institute (KEMRI), Nairobi, Kenya (SERU #4035). Individual patient consent was not required by the committee for the use of these samples for genomic surveillance to inform public health response.

### Study period and population

This study examined samples collected between 17^th^ March 2020 to 26^th^ February 2021 from six counties in Coastal Kenya i.e. Mombasa, Kilifi, Kwale, Tana River, Taita Taveta and Lamu. We divided the study period into three phases based on trends in daily reported COVID-19 national case numbers by the MoH (MOH, 2021): (a) Introduction phase – 17^th^ March-20^th^ May, 2020 (b) Wave one – 21^st^ May-15^th^ September 2020 and (c) Wave two – 16^th^ September to 28^th^ February 2021, **Figure 1**. We considered the transition from introduction phase to Wave one as the timepoint when the national daily positives exceeded 50 and the transition from Wave one to Wave two as the timepoint when a consistent renewed rise of national daily number of positives started after Wave one peak.

### Patient samples

This study analyzed SARS-CoV-2 positive NP/OP swab samples that were collected by the MoH County Department of Health RRTs, across all the six counties of Coastal Kenya. SARS-CoV-2 positive diagnosis was made at the KWTRP in Kilifi County (Agoti et al., 2021). The RRTs delivered the NP/OP swabs to KWTRP laboratories within 48 hours of collection in cool boxes with ice packs. The samples were from persons of any age and sampling followed the MOH eligibility criteria that were revised from time to time (Githinji et al., 2020). Persons sampled included those with acute respiratory symptoms, those with a recent history of travel to the early COVID-19 hotspots, contacts of confirmed cases, persons presenting at international border points seeking entry into Kenya, among other criteria.

### SARS-CoV-2 detection at KWTRP

Viral RNA was extracted from the NP/OP samples using any one of seven commercial kits that were available namely, QIAamp Viral RNA Mini Kit, RNeasy ® QIAcube ® HT Kit, QIASYMPHONY ® RNA Kit, T0IANamp Virus RNA Kit, Da An Gene Nucleic acid Isolation and Purification Kit, SPIN X Extraction and RADI COVID-19 detection Kit. The extracts were tested for the presence of SARS-CoV-2 nucleic acid following different protocols depending on which one was available of the following 7 kits/protocols: 1) the Berlin (Charité) primer-probe set (targeting envelope (E) gene, nucleocapsid (N) or RNA-dependent RNA-polymerase (RdRp)), 2) European Virus Archive – GLOBAL (EVA-g) (targeting E or RdRp genes), 3) Da An Gene Co. detection Kit (targeting N or ORF1ab), 4) BGI RT-PCR kit (targeting ORF1ab), 5) Sansure Biotech Novel Coronavirus (2019-nCoV) Nucleic Acid Diagnostic real-time RT-PCR kit or 6) Standard M kit (targeting E and ORF1ab) and 7) TIB MOLBIOL kit (targeting E gene). Protocol-specific recommended cycle threshold cut-offs were followed in defining SARS-CoV-2 positives.

### SARS-CoV-2 genome sequencing

Only samples that had a RT-PCR cycle threshold value of <30.0 were targeted for whole genome sequencing (Githinji et al., 2020), **Supplementary-Figure 1**. Viral RNA from positive samples was reextracted using QIAamp Viral RNA Mini kit following the manufacturer’s instructions and reverse transcribed using LunaScript® RT SuperMix Kit. The cDNA was amplified using Q5® Hot Start High-Fidelity 2x Mastermix along with the ARTIC nCoV-2019 version 3 primers. The PCR products were run on a 1.5% agarose gel and for samples whose SARS-CoV-2 amplification was considered successful were purified using Agencourt AMPure XP beads and taken forward for library preparation.

Sequencing libraries were constructed using Oxford Nanopore Technology (ONT) ligation sequencing kit and the ONT Native Barcoding Expansion kit as described in the ARTIC protocol (Tyson et al., 2020). Every MinION (Mk1B) run comprised 23 samples and one negative (no-template) control.

### SARS-CoV-2 genome assembly

Following MinION sequencing, the FAST5 files were base-called and demultiplexed using the OTN’s software Guppy v3.5-4.2. For base calling, high accuracy mode was used. Consensus SARS-CoV-2 sequences were derived from the reads using the ARTIC bioinformatics pipeline (https://artic.network/ncov-2019/ncov2019-bioinformatics-sop.html; last accessed 2021-08-03). A threshold of ×20 read depth was required for a base to be included in the consensus genome otherwise it was masked with an N. Only complete or near-complete genomes N count < 5,980 (i.e >80% coverage) were taken forward for phylogenetic analysis.

### Lineage assignment and calling of amino acid changes

The consensus genomes were assigned into lineages using the PANGOLIN nomenclature (Rambaut et al., 2020). We used the command line pangolin v3.1.10 software to assign Pango lineages to each consensus sequence with pango v1.2.46 and pangoLEARN model v2021-07-28 (O’Toole et al., 2021). Contextual information about lineages was obtained from the Lineage list available at https://cov-lineages.org/lineage_list.html (last accessed 2021-08-04). Variants of concern (VOC) and variants of interest (VOI) were designated based on the WHO framework as of 31^st^ May 2021 (https://www.who.int/en/activities/tracking-SARS-CoV-2-variants/).

The amino acid changes in the Coastal Kenya genomes from the reference strain Wuhan-Hu-1/2019 was investigated using the nextclade tool v0.14.2 (Hadfield et al., 2018): https://clades.nextstrain.org/; last accessed 2021-08-03. Mutations in the Kenyan lineages were visualized using the tool available on Stanford University CORONAVIRUS ANTIVIRAL & RESISTANCE Database webpage: https://covdb.stanford.edu/page/mutation-viewer/; last accessed 2021-08-03).

### Global contextual sequences

We prepared three sets of contextual sequences from GISAID database (https://www.gisaid.org/). Selected sequences were those with non-ambiguous sampling date, sampled during the period covered by our study (March 2020 – February 2021) and after alignment possessed <5,980 ambiguous (N) nucleotides i.e. showed >80% genome completeness.

a. **Set 1:** We compiled an African SARS-CoV-2 genome set which we assigned Pango lineages and used to compare the continental lineage temporal patterns to the Coastal Kenya lineage distribution. A total of 12,191 genomes were included for this purpose downloaded on the 11^th^ of May 2021. From these we also created an Eastern Africa subset which comprised of 2,261 genomes from 10 countries, namely, Zimbabwe, Zambia, Uganda, Rwanda, Reunion (a France overseas territory), Mozambique, Malawi, Madagascar, Ethiopia and Comoros.
b. **Set 2**: We compiled a global reference set of 24,511 genomes collected across all the six inhabited continents between 17^th^ March 2020 and 27^th^ February 2021. The genomes were selected from all available genomes in GISAID database using the R randomization command: *sample_n()*. Between 1916 and 2174 genomes were selected for each included month (mean of 2042 and median of 2049) randomly selected across all continents from across 164 countries. These genomes were used to infer the global temporal patterns of the lineages observed in Coastal Kenya. A subset of these genomes (n=4000) was combined with the Kenyan genomes to infer their global phylogenetic context and in the import/export analysis described below.
c. **Set 3**: We compiled a genotype reference for the top six observed lineages in Coastal Kenya. For lineages B.1. and B.1.1 which >5000 genomes exist in GISAID were prepared we used the sub-sample from Set 2 above. For lineages B.1.530, B.1.549, B.1.596.1 and N.8 we retrieved all genomes assigned these lineages available from GISAID to infer their global phylogenetic context.

### Phylogenetic analysis

Multiple sequence alignments were prepared in Nextalign v 0.1.6 software and using the initial Wuhan sequence (Accession number: NC_045512) as the reference, command:

*nextalign -r NC_045512*.*fasta -i input*.*fasta*

The alignment was manually inspected in AliView v1.21 to spot any obvious misalignments. Quick non-bootstrapped neighbor joining trees were created in SEAVIEW v4.6.4 to identify any aberrant sequences that were henceforth discarded. We reconstructed maximum likelihood (ML) phylogenies using IQTREE v2.1.3, command:

.*/iqtree2 -s input*.*aligned*.*fasta -nt 4*

The software initiates tree reconstruction after assessment and selection of the best model of nucleotide substitution for the alignment using ModelFinder. For example for the large ML phylogeny presented in Figure 5A, the Best-fit mode was determined as GTR+F+R3.

The ML trees were linked to the various metadata (lineage, county, source etc) in R programming software v4.0.2 and visualized using R ggTree v2.4.2. TempEst v1.5.3 was used to assess the presence of a molecular clock signal in analysed data and linear regression of root-to-tip genetic distances against sampling dates plotted in RStudio and the correlation coefficient assessed.

### Import/export analysis

The global ML tree topology (**Figure 5**) was used to estimate the number of viral transmission events between Coastal Kenya and the rest of the world as in (Wilkinson et al.). The software TreeTime was used to transform the ML tree topology into a dated tree assuming a constant genomic evolutionary rate of SARS-CoV-2 of 8.4 ×10^−4^ nucleotide substitutions per site per year (Sagulenko et al., 2018), command:

*treetime --tre input*.*aligned*.*fasta*.*treefile --aln input*.*aligned*.*fasta*

*-–clock-rate 0*.*00084 –-dates dates*.*csv*

Outlier sequences were identified by TreeTime and excluded during this process. A migration model was henceforth fitted to the resulting time-scaled phylogenetic tree from TreeTime, mapping the location status of the genomes from the six counties at both the tips and internal nodes, command:

*treetime mugration –-tree input*.*tree*.*nwk –-states mugration*.*csv –-attribute Location*

Using the date and location annotated tree topology, we counted the number of transitions between and within Coastal Kenya counties and the rest of the world using the python script developed and described by the team at KRISP (https://github.com/krisp-kwazulu-natal/SARSCoV2_South_Africa_major_lineages/tree/main/Phylogenetics; last accessed 2020-08-04) and plotted this using ggplot2 v3.3.3. This analysis was repeated with different sub-sample of the global background data and with a down sample of the coastal Kenya genomes that was normalized spatial-temporally (using a maximum of 15 samples per county per month).

### Epidemiological data

The Kenya daily case data for the period between March 2020 and February 2021 was downloaded from Our World in Data database (https://ourworldindata.org/coronavirus/country/kenya). Metadata for the Coastal Kenya samples was compiled from the MoH case investigation forms delivered alongside the samples.

### Kenya COVID-19 response

We derived status of Kenya government COVID-19 interventions using data available from Our World in Data database that has calculated the Oxford Stringency Index (SI). These SI estimations are composite measure based on nine response indicators implemented by governments rescaled to a value 0-100, with 100 being strictest (Hale et al., Noam Angrist). The indicators are: (a) School closures, (b) Workplace closures, (c) Cancellation of public events, (d) Restrictions on public gatherings, (e) Closures of public transport, (f) Stay-at-home requirements, (g) Public information campaigns, (h) Restrictions on internal movements and (d) International travel controls. These are The Kenya government revised the policy interventions at approximately monthly intervals (Brand et al., 2021) and the changes in SI overtime are shown in **Figure 1D**.

### Statistical analysis

Statistical data analyses were performed in R v 4.0.5. Summary statistics including proportions, means, median and ranges were calculated and provided where applicable. T-test was used to evaluate a statistical difference in means of the pairwise nucleotide or amino acid differences between the coastal and global sample. The *lm* function in R was used to fit a linear regression model evaluating the relationship between sampling dates and root-to-tip genetic distance in the ML phylogeny. The goodness of fit was inferred from the correlation coefficient.

## Data Availability

Coastal Kenya genomes have been deposited to both GISAID (Accession numbers: EPI_ISL_1039223-29; EPI_ISL1440075-109, IPI_ISL_457845-931; EPI_ISL_568695-872; EPI_ISL806548-717; EPI_ISL_855494-548; EPI_ISL_968807-9245).

## Data availability

The data and scripts used to generate the figures shown in the manuscript are hosted on Harvard dataverse: DOI: https://doi.org/10.7910/DVN/4ZZYIM.

The Coastal Kenya genomes are available in the GISAID database (Accession numbers: EPI_ISL_1039223-29; EPI_ISL1440075-109, IPI_ISL_457845-931; EPI_ISL_568695-872; EPI_ISL806548-717; EPI_ISL_855494-548; EPI_ISL_968807-9245).

## Acknowledgements

We thank (a) the members of the six Coastal counties of Kenya RRTs for collecting the samples analysed here; (b) the members of the COVID-19 KWTRP Testing Team who tirelessly analysed the samples received at KWTRP to identify positives (see full list of members below); (c) the KWTRP data entry team, (d) Laboratories that have shared sequence data on GISAID that we included as comparison data in our analysis (see list in appendix); (e) the KRISP team in South Africa for sharing the scripts we used in the import/export analysis and AFRICA-CDC for facilitating Africa genomics training. This paper is published with permission of the Director of KEMRI.

## Members of COVID-19 Testing Team at KWTRP

Agnes Mutiso, Alfred Mwanzu, Angela Karani, Bonface M. Gichuki, Boniface Karia, Brian Bartilol, Brian Tawa, Calleb Odundo, Caroline Ngetsa, Clement Lewa, Daisy Mugo, David Amadi, David Ireri, Debra Riako, Domtila Kimani, Edwin Machanja, Elijah Gicherua, Elisha Omer, Faith Gambo, Horace Gumba, Isaac Musungu, James Chemweno, Janet Thoya, Jedida Mwacharo, John Gitonga, Johnstone Makale, Justine Getonto, Kelly Ominde, Kelvias Keter, Lydia Nyamako, Margaret Nunah, Martin Mutunga, Metrine Tendwa, Moses Mosobo, Nelson Ouma, Nicole Achieng, Patience Kiyuka, Perpetual Wanjiku, Peter Mwaura, Rita Warui, Robinson Cheruiyot, Salim Mwarumba, Shaban Mwangi, Shadrack Mutua, Sharon Owuor, Susan Njuguna, Victor Osoti, Wesley Cheruiyot, Wilfred Nyamu, Wilson Gumbi and Yiakon Sein.

## Supplementary Material for

### Additional available material relating to this manuscript

STROBE checklist

Transparent Reporting File

Source code for figures presented in the manuscript

Accession numbers of coastal Kenya genomes presented in this study

Acknowledgements GISAID depositors

**Supplementary-Figure 1.**
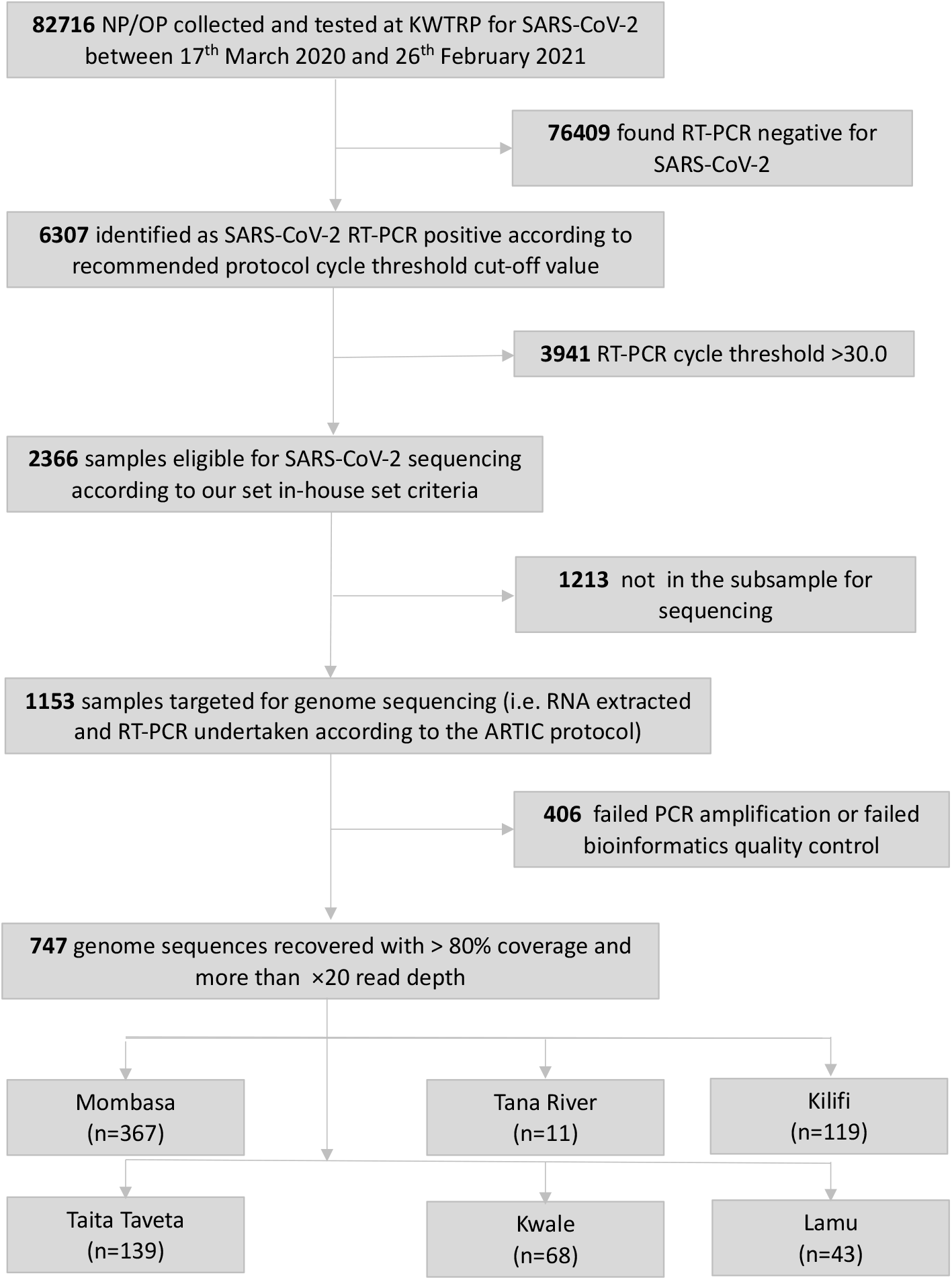
Flow of samples analysed in this study.

**Supplementary-Figure 2.**
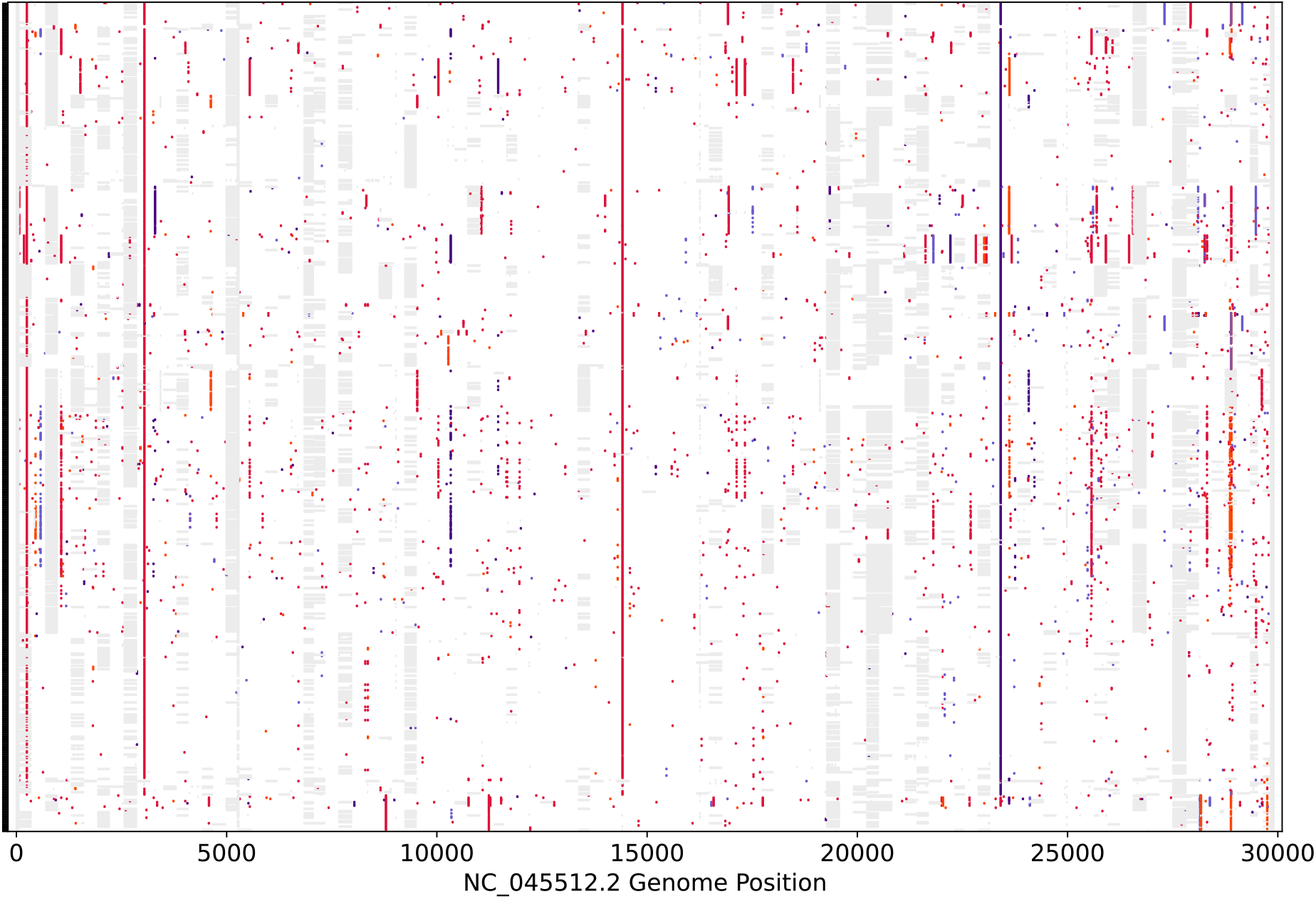
Nucleotide differences in SARS-CoV-2 genome sequences from the coastal Kenya samples compared to the reference genome Wuhan-Hu-1 (Accession #: NC_045512.2). Vertical coloured bars show the nucleotide differences from the reference genome; orange is a change to A, crimson is a change to T, indigo a change to G, and slate blue a change to C. Grey is a nucleotide deletion or an area not sequenced in our labouratory due to amplicon drop-off or inconclusive assembly.

**Supplementary-Figure 3.**
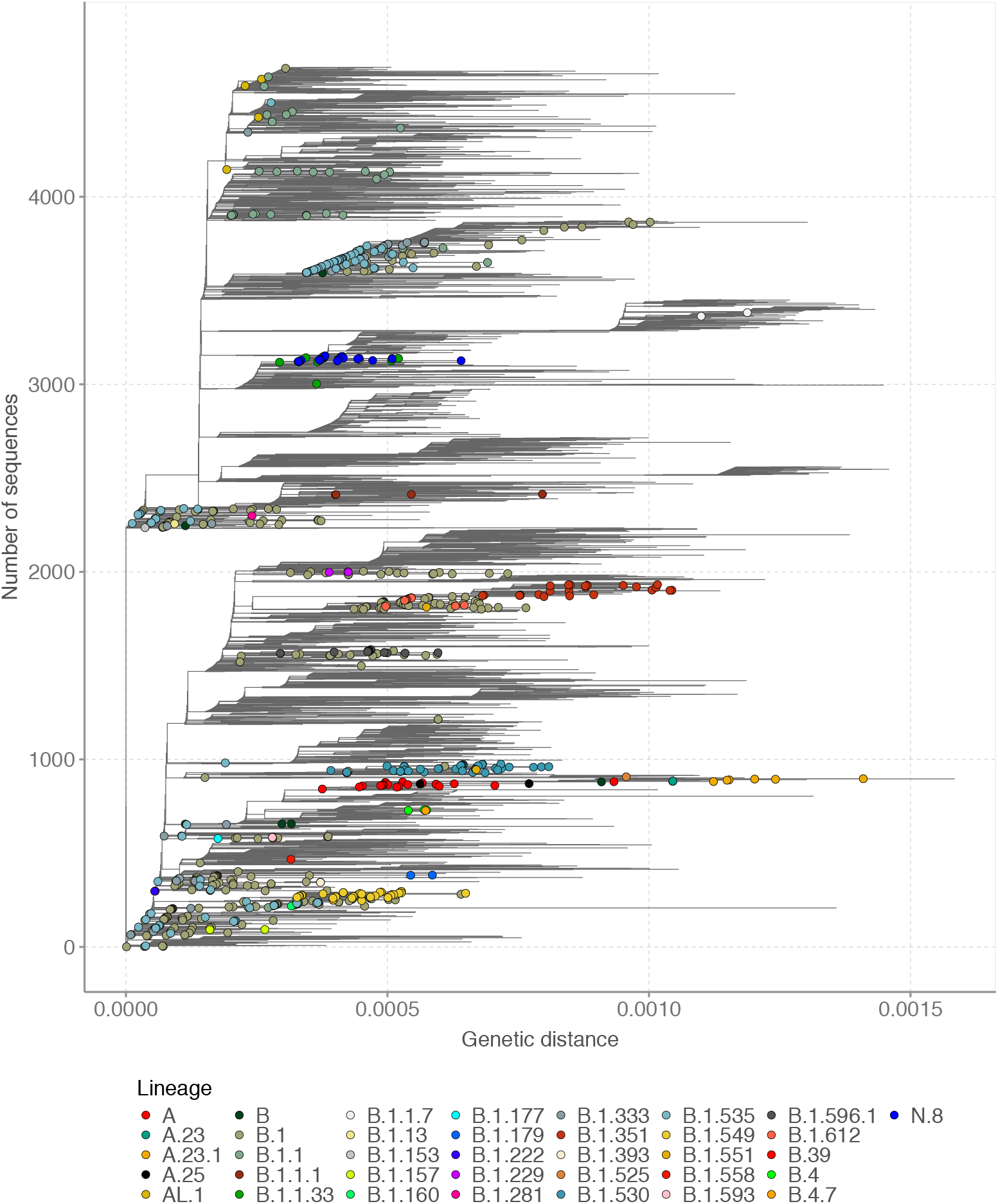
Mutation-resolved global phylogenetic tree showing the genetic closeness of the sequences from coastal Kenya while including a representative global sub-sample. The tree uses the same data as Figure 5A in the main test (time-resolved tree) by combining 3,958 global sequences and 732 coastal Kenya sequences. The coastal Kenya sequences are indicated by a filled circle symbol, coloured by the assigned Pango lineage.

**Supplementary-Figure 4.**
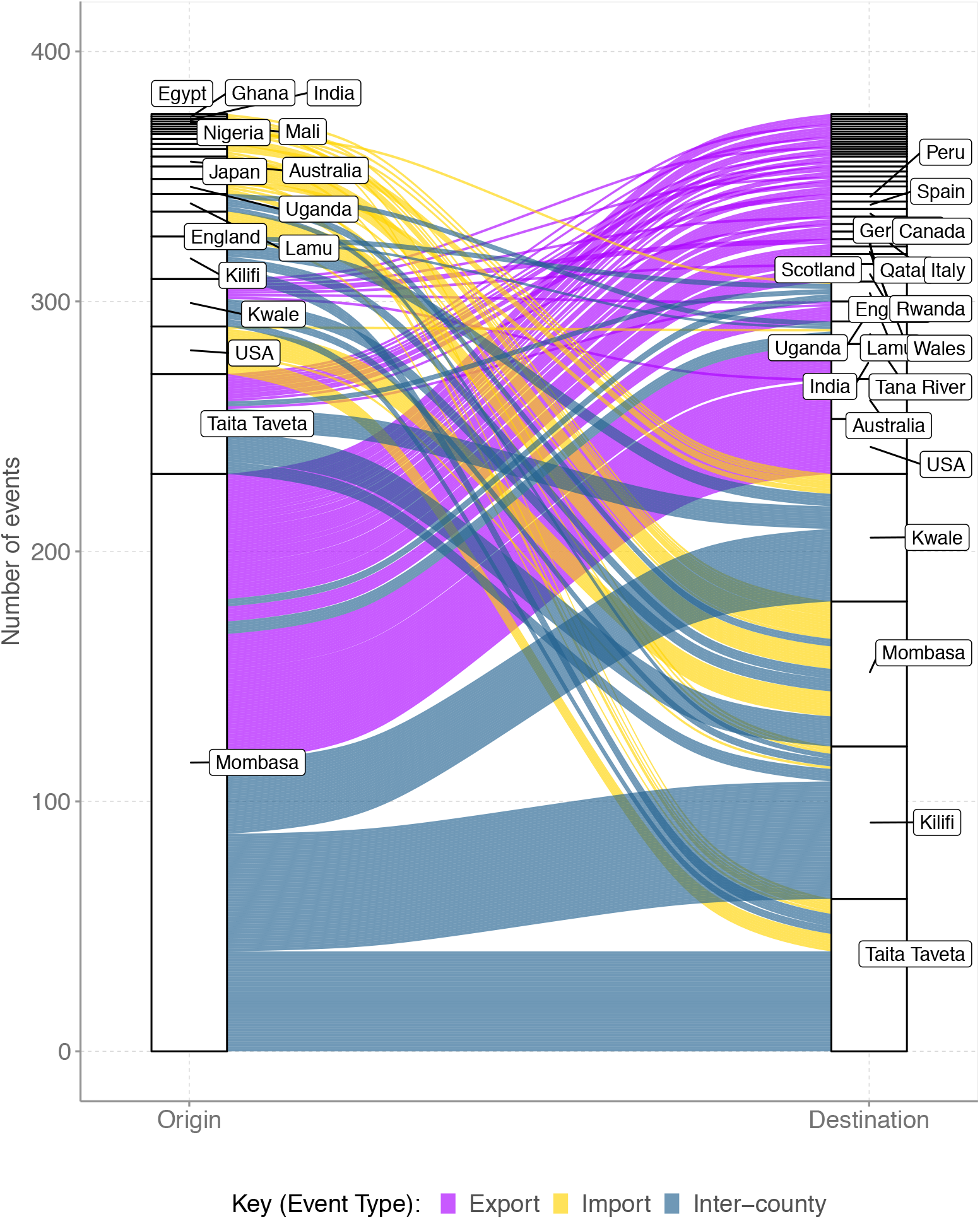
An alluvium plot showing the number of viral imports and exports from coastal Kenya over the study period. This figure, unlike Figure 8A, provides the details of the countries from which import to or exports from coastal Kenya occurred. Exports from coastal Kenya are depicted by a purple colour, imports to costal Kenya are depicted by yellow colour, within coastal Kenya intercounty spread is indicated by the cadet blue colour.

**Supplementary-Figure 5.**
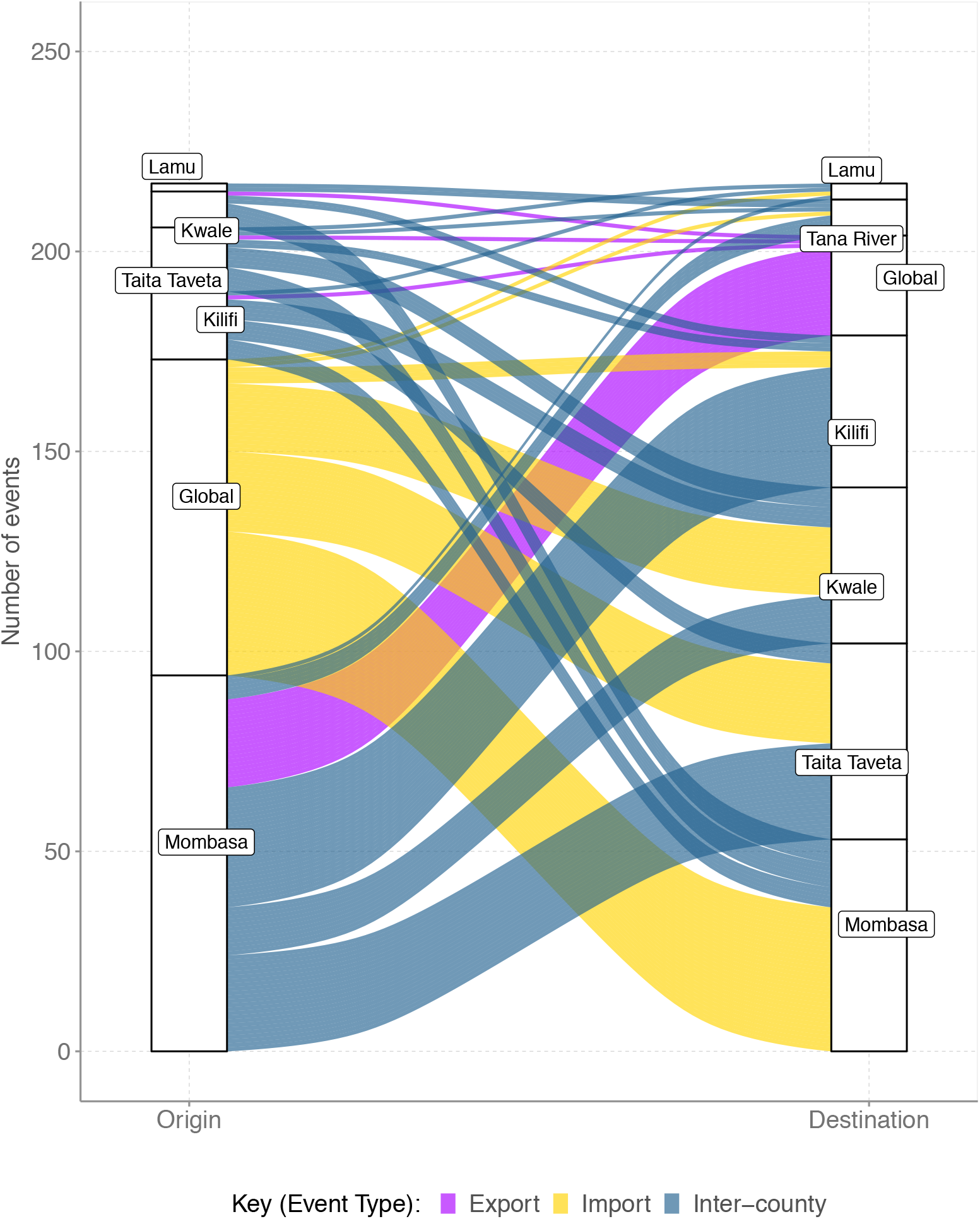
An alluvium plot showing the number of viral imports and exports from coastal Kenya over the study period using a subsample of the coastal Kenya genomes (n=360). The sub-sample was randomly selected aiming to be spatial-temporally representative with up to 15 genomes per county per month over the study period. Exports are depicted by purple colour, imports are depicted by yellow colour, within coastal Kenya intercounty spread is indicated by the cadet blue colour.

**Supplementary-Figure 6.**
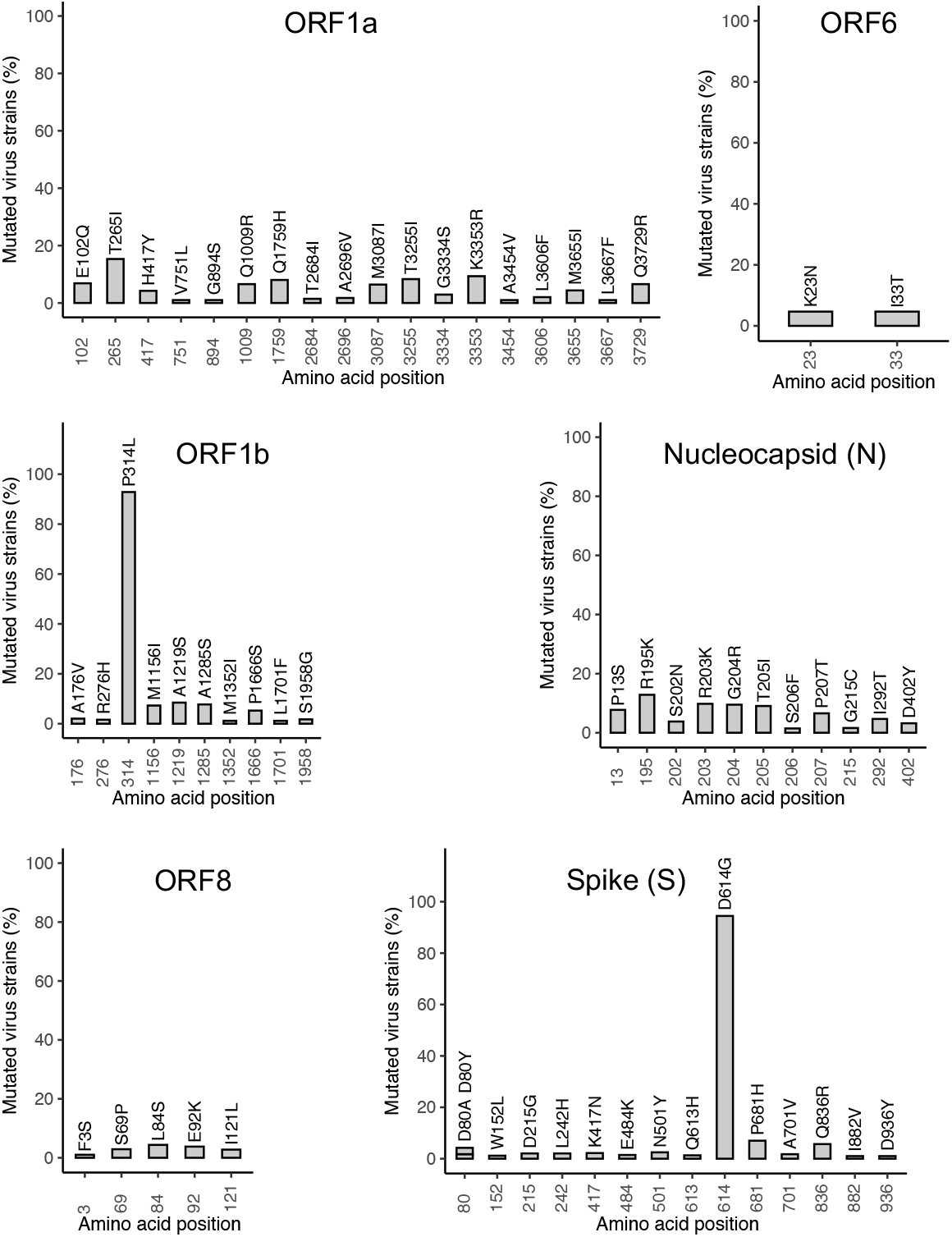
Position of non-synonymous mutations in the 747 genomes from coastal Kenya compare with the Wuhan-Hu-1 reference genome (Accession number NC_045512). Only changes occurring in >1% of genomes are shown. The percentage frequency of the changes in the 747 genomes is shown on the y-axis.

**Supplementary-Figure 7.**
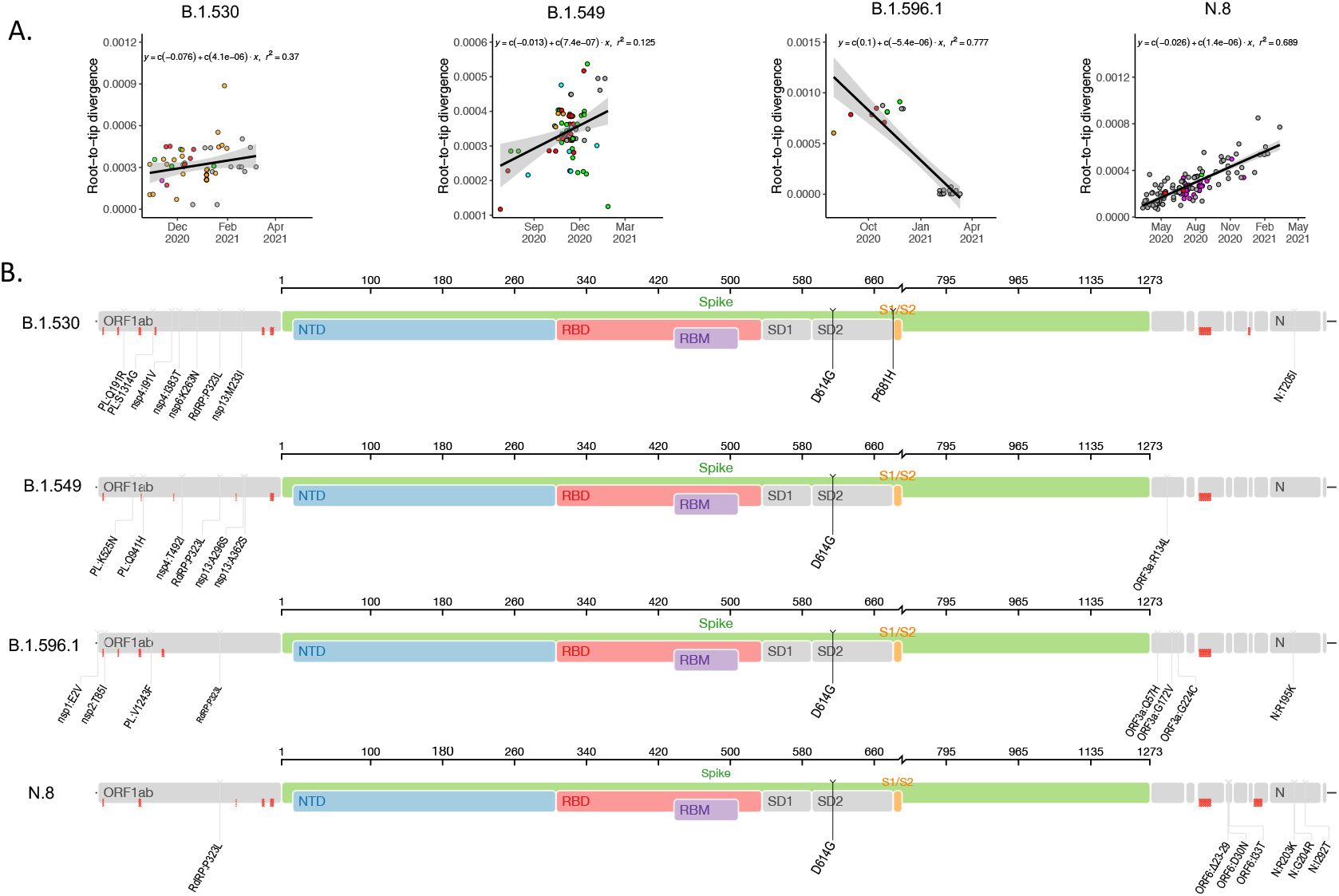
Genomic variation characteristics of the four lineages first identified in Kenya. Panel A, root-to-tip regression plots for the four lineages first identified in Kenya. Panel B, Genome maps of the “Kenya” lineages where the spike region is shown in detail and in colour and the rest of the genome is shown in grey colour. Red marks indicate where sequencing of Kenya strains resulted in ambiguous nucleotides. Plots generated using tool available from Stanford University Web-page Coronavirus antiviral and resistance database https://covdb.stanford.edu/page/mutation-viewer/: last accessed on 2021-08-03.

**Supplementary-Table 1.** Summary output from separate runs of the import/export ancestral state reconstruction (ASR) analysis

| **Characteristic** | **Run 1<sup>#</sup>** | **Run 2<sup>£</sup>** | **Run 3<sup>§</sup>** |
| --- | --- | --- | --- |
| **Total Location transition events** | 357 | 341 | 217 |
| **International imports** |  |  |  |
| **Total** | 69 | 65 | 79 |
| **Through Mombasa** | 40 | 38 | 36 |
| **Through Kwale** | 9 | 5 | 17 |
| **Through Taita Taveta** | 13 | 13 | 20 |
| **Through Kilifi** | 5 | 5 | 17 |
| **Through Lamu** | 1 | 1 | 1 |
| **International exports** |  |  |  |
| **Total** | 93 | 87 | 25 |
| **Through Mombasa** | 86 | 81 | 22 |
| **Through Kwale** | 6 | 3 | 1 |
| **Through Taita Taveta** | 1 | 3 | 1 |
| **Through Kilifi** | - | - | 1 |
| **Inter-county transmission events** | 193 | 189 | 113 |

<sup>#</sup>Import/export analysis results from ASR combining 732 genome sequences from coastal Kenya that passed the molecular clock quality control and a random 3,958 global (outside Kenya) genomes.
<sup>£</sup>Import/export analysis results from ASR combining 728 genomes from coastal Kenya passing the molecular clock quality control and 3,968 global (outside Kenya) genomes.
<sup>§</sup> Import/export analysis results from ASR combining 338 genome sequences from coastal Kenya that passed the molecular clock quality control and a random 3979 global (outside Kenya) genomes. The coastal Kenya sub-sample used for this run 3 was randomly selected aiming to be spatial-temporally representative with up to 15 genomes per county per month over the study period.

